# Subcutaneous REGEN-COV Antibody Combination in Early Asymptomatic SARS-CoV-2 Infection: A Randomized Clinical Trial

**DOI:** 10.1101/2021.06.14.21258569

**Authors:** Meagan P. O’Brien, Eduardo Forleo-Neto, Neena Sarkar, Flonza Isa, Peijie Hou, Kuo-Chen Chan, Bret J. Musser, Katharine J. Bar, Ruanne V. Barnabas, Dan H. Barouch, Myron S. Cohen, Christopher B. Hurt, Dale R. Burwen, Mary A. Marovich, Elizabeth R. Brown, Ingeborg Heirman, John D. Davis, Kenneth C. Turner, Divya Ramesh, Adnan Mahmood, Andrea T. Hooper, Jennifer D. Hamilton, Yunji Kim, Lisa A. Purcell, Alina Baum, Christos A. Kyratsous, James Krainson, Richard Perez-Perez, Rizwana Mohseni, Bari Kowal, A. Thomas DiCioccio, Neil Stahl, Leah Lipsich, Ned Braunstein, Gary Herman, George D. Yancopoulos, David M. Weinreich, for the COVID-19 Phase 3 Prevention Trial Team

## Abstract

**Importance:** Easy-to-administer antiviral treatments may be used to prevent progression from asymptomatic infection to COVID-19 and to reduce viral carriage.

**Objective:** Evaluate the efficacy and safety of subcutaneous casirivimab and imdevimab antibody combination (REGEN-COV) to prevent progression from early asymptomatic SARS-CoV-2 infection to COVID-19.

**Design:** Randomized, double-blind, placebo-controlled, phase 3 study that enrolled asymptomatic close contacts living with a SARS-CoV-2–infected household member (index case). Participants who were SARS-CoV-2 RT-qPCR–positive at baseline were included in the analysis reported here.

**Setting:** Multicenter trial conducted at 112 sites in the United States, Romania, and Moldova.

**Participants:** Asymptomatic individuals ≥12 years of age were eligible if identified within 96 hours of collection of the index case’s positive SARS-CoV-2 test sample.

**Interventions:** A total of 314 asymptomatic, SARS-CoV-2 RT-qPCR–positive individuals living with an infected household contact were randomized 1:1 to receive a single dose of subcutaneous REGEN-COV 1200mg (n=158) or placebo (n=156).

**Main Outcome(s) and Measure(s):** The primary endpoint was the proportion of participants who developed symptomatic COVID-19 during the 28-day efficacy assessment period. The key secondary efficacy endpoints were the number of weeks of symptomatic SARS-CoV-2 infection and the number of weeks of high viral load (>4 log_10_ copies/mL). Safety was assessed in all treated participants.

**Results:** Subcutaneous REGEN-COV 1200mg significantly prevented progression from asymptomatic to symptomatic disease compared with placebo (31.5% relative risk reduction; 29/100 [29.0%] vs 44/104 [42.3%], respectively; *P*=.0380). REGEN-COV reduced the overall population burden of high–viral load weeks (39.7% reduction vs placebo; 48 vs 82 total weeks; *P*=.0010) and of symptomatic weeks (45.3% reduction vs placebo; 89.6 vs 170.3 total weeks; *P*=.0273), the latter corresponding to an approximately 5.6-day reduction in symptom duration per symptomatic participant. Six placebo-treated participants had a COVID-19–related hospitalization or ER visit versus none for those receiving REGEN-COV. The proportion of participants receiving placebo who had ≥1 treatment-emergent adverse events was 48.1% compared with 33.5% for those receiving REGEN-COV, including events related (39.7% vs 25.8%, respectively) or not related (16.0% vs 11.0%, respectively) to COVID-19.

**Conclusions and Relevance:** Subcutaneous REGEN-COV 1200mg prevented progression from asymptomatic SARS-CoV-2 infection to COVID-19, reduced the duration of high viral load and symptoms, and was well tolerated.

**Trial Registration:** ClinicalTrials.gov Identifier, NCT04452318

**KEY POINTS:** *Question:* Can treatment with the anti–SARS-CoV-2 antibody combination REGEN-COV prevent COVID-19 and reduce viral load when given to recently exposed and asymptomatic individuals?

*Findings:* In this randomized, double-blind, phase 3 trial, subcutaneously administered REGEN-COV 1200 mg significantly reduced progression of asymptomatic SARS-CoV-2 infection to symptomatic infection (ie, COVID-19) by 31.5% compared with placebo. REGEN-COV also reduced the overall population burden of high viral load weeks (39.7% reduction vs placebo; 48 vs 82 total weeks; *P*=.0010).

*Meaning:* In the current pandemic, utilization of subcutaneous REGEN-COV prevents progression of early asymptomatic infection to COVID-19 and reduces viral carriage.

## INTRODUCTION

SARS-CoV-2, a betacoronavirus first identified in China at the end of 2019, is the cause of COVID-19 and the current global pandemic that has infected over 200 million and resulted in the deaths of over 4 million persons worldwide as of July 2021.^1,2^ Control of SARS-CoV-2 and the pandemic has been challenging, due in part to a highly variable incubation period (range 2–14 days) and high rates of transmission from asymptomatic individuals.^3-8^ Moreover, numerous SARS-CoV-2 variants of concern/interest (VOCs/VOIs) have emerged and are currently circulating globally, eg, B.1.1.7 (alpha), B.1.351 (beta), P.1 (gamma), B.1.617.2 (delta), and B.1.427/429 (epsilon).^9,10^ As of July 2021, VOC delta, which has enhanced transmissibility, has become the dominant strain worldwide.^10-12^ Even with the arrival of effective SARS-CoV-2 vaccines,^13,14^ many individuals across the globe remain at risk of infection and potential progression to severe COVID-19 or longer-term COVID-19–related complications due to multiple factors, including limited vaccine uptake in certain geographic areas, inadequate responses to vaccination in certain risk groups, or immune-evading VOCs.^1,15-18^ For those individuals not protected by vaccination, complementary approaches for immediate protection are needed.

REGEN-COV consists of two neutralizing monoclonal antibodies, casirivimab and imdevimab, that bind distinct, non-overlapping epitopes on the SARS-CoV-2 spike protein receptor binding domain and block virus entry.^19^ The two-antibody combination reduces the risk of emergence of treatment-induced SARS-CoV-2 variants and retains neutralization potency in vitro against already-circulating variants of concern or interest (VOC/VOIs), including B.1.1.7 (alpha), B.1.351 (beta), P.1 (gamma), B.1.617 (delta), and B.1.427/429.^20-23^ REGEN-COV has proven effective in treating COVID-19 outpatients^24,25^ and is currently authorized in the U.S. under an emergency use authorization (EUA) for the treatment of mild-to-moderate COVID-19 and for post-exposure prophylaxis in certain individuals exposed to or at high risk of exposure to a SARS-CoV-2–infected individual.^26,27^

We conducted a phase 3, randomized controlled trial in asymptomatic household contacts of SARS-CoV-2–infected individuals to assess whether subcutaneous REGEN-COV 1200 mg (600 mg of each monoclonal antibody) may prevent infection and/or COVID-19 in this high-risk setting. Nasopharyngeal swabs for SARS-CoV-2 RT-qPCR were performed prior to randomization and were used to assign participants into 2 analysis cohorts referred to as Part A and B: Part A for SARS-CoV-2 RT-qPCR– negative participants and Part B for SARS-CoV-2 RT-qPCR–positive participants. We recently reported that REGEN-COV significantly prevented symptomatic SARS-CoV-2 infection compared with placebo (81.4% risk reduction) in uninfected close contacts (Part A).^28^ Here, we describe the results from asymptomatic, infected close contacts (Part B) treated with subcutaneous REGEN-COV 1200 mg.

## METHODS

### Trial Design

This randomized, double-blind, placebo-controlled, phase 3 trial assessed the efficacy and safety of REGEN-COV to prevent the progression to symptomatic SARS-CoV-2 infection among asymptomatic, infected household contacts of infected individuals (ClinicalTrials.gov number, NCT04452318). The trial was conducted at 112 sites in the United States, Romania, and Moldova and was managed jointly by Regeneron, the COVID-19 Prevention Network (CoVPN), and the National Institute of Allergy and Infectious Diseases (NIAID).

Nasopharyngeal and serum samples were collected at the screening/baseline visit for central lab RT-qPCR testing and serum antibody testing. RT-qPCR was used to determine whether the study participant was infected with SARS-CoV-2, while baseline serology testing for serum anti–SARS-CoV-2 antibodies (anti-spike [S1] IgA, anti-spike [S1] IgG, and anti-nucleocapsid IgG) was used to determine if the participant had evidence of immunity to SARS-CoV-2 (ie, seropositive, as opposed to seronegative). Part A analyses included those who at baseline were asymptomatic and RT-qPCR– negative; Part B analyses included those who at baseline were asymptomatic and RT-qPCR–positive (**Figure S1**). Here, we describe the primary results for Part B.

Study participants were randomized (1:1) to receive REGEN-COV or placebo and stratified by SARS-CoV-2 local diagnostic results, when available, and age (see **Supplementary Appendix**). The trial consisted of a 1-day screening/baseline period, a 28-day efficacy assessment period (EAP), and a 7-month follow-up period (**Figure S1**). **Protocols** (original and final/Amendment 6) for this primary analysis and a summary of protocol amendments are included in the **Supplement**.

### Trial Oversight

The trial was conducted in accordance with the principles of the Declaration of Helsinki, GCP/ICH-E-9 guidelines, and all applicable regulatory standards. All participants provided written informed consent. Additional details are provided in the **Supplementary Appendix**.

### Study Participants

The study included adults (≥18 years of age) and adolescents (≥12 to <18 years of age) who were household contacts of the first known household member with SARS-CoV-2 infection (index case) and asymptomatic (having no active respiratory or non-respiratory symptoms consistent with COVID-19). Part B participants were SARS-CoV-2–positive by RT-qPCR but unaware of being infected. Individuals were eligible for inclusion in the study regardless of serum antibody status (seronegative/seropositive/sero-undetermined). Randomization occurred within 96 hours of collection of the index case’s positive SARS-CoV-2 test sample. The full list of inclusion/exclusion criteria are provided in the **Supplementary Appendix**.

### Intervention and Assessments

At baseline (day 1), participants received a single dose of REGEN-COV 1200 mg (120 mg/mL) or placebo via subcutaneous (SC) injection.

Signs and symptoms of COVID-19 were collected by weekly investigator-led interviews. At each visit/contact, the investigator interviewed the participant about adverse events they were experiencing or could have experienced since the last visit/contact. If the participant developed signs and/or symptoms, this data was collected weekly until resolved.

Serial NP swabs were collected at baseline, prior to study drug administration, and weekly during the EAP and/or the follow-up period to determine SARS-CoV-2 viral load by RT-qPCR until participants tested negative on two consecutive swabs.

Details on the intervention, assessments, and analytical methods are provided in the **Protocol** or **Supplementary Appendix** or have been previously described.^25^

### Endpoints

The primary efficacy endpoint was the proportion of participants who developed signs and symptoms of COVID-19 within 14 days of a positive RT-qPCR at baseline or during the 28-day efficacy assessment period. A broad definition (broad term) of what constituted symptomatic COVID-19 was used for the primary analysis; alternative definitions (strict-term and CDC) were used for other analyses (see **Supplementary Appendix**).

The key secondary efficacy endpoints were the number of weeks (in the overall population) of symptomatic SARS-CoV-2 infection (broad-term) and the number of weeks of high viral load (>4 log_10_ copies/mL) in nasopharyngeal samples over 28 days. The full lists of secondary efficacy and exploratory endpoints and methods of calculation are provided in the **Statistical Analysis Plan** or **Supplementary Appendix**.

Safety endpoints included the collection of serious adverse events (SAEs), treatment-emergent adverse events (TEAEs), and adverse events of special interest (AESIs) defined as grade ≥3 injection site reactions or hypersensitivity reactions.

### Statistical Analysis

The statistical analysis plan for the presented analysis was finalized prior to database lock and treatment unblinding. The primary analysis population used for efficacy analyses was the seronegative modified full analysis set (seronegative mFAS-B): all randomized participants who were asymptomatic, negative for SARS-CoV-2 antibodies, and SARS-CoV-2–positive by central lab RT-qPCR at baseline (ie, part B in this two-part trial). Additional secondary and exploratory efficacy analyses were performed on data from participants regardless of baseline serostatus. Efficacy analyses are reported through the 28-day EAP, unless otherwise noted. The safety analysis set for part B (SAF-B) included all participants who received study drug (active or placebo). Efficacy and safety analyses are reported for participants randomized through January 28, 2021, until the data cut-off date of March 11, 2021.

The sample size calculation assumed that approximately 200 seronegative participants would be enrolled and 50% of placebo participants would develop symptoms; this would provide >90% power to detect a relative risk of 0.5 at a two-sided alpha level of 0.05.

Hierarchical testing was employed for the primary and key secondary endpoints to control for type 1 error based on a two-sided alpha of 0.05. Participants with COVID-19 symptoms who were missing central lab RT-qPCR test results were considered as having a symptomatic infection if any symptoms occurred within 14 days of a positive SARS-CoV-2 local test.

Additional statistical methods are described in more detail in the **Statistical Analysis Plan** in the **Supplement**.

## RESULTS

### Trial Population

Between July 13, 2020 and January 28, 2021, a total of 314 household contacts were confirmed SARS-CoV-2 RT-qPCR–positive (PCR-positive) at baseline based on central lab nasopharyngeal RT-qPCR; 156 study participants received placebo, and 155 received REGEN-COV 1200 mg SC (3 randomized participants did not receive any study drug).

The primary and key secondary analyses were performed in participants without evidence of immunity (ie, seronegative); however, all participants, regardless of serostatus were included in other secondary and exploratory analyses. Of the 314 PCR-positive participants randomized, 207 (66%) were seronegative, 84 were seropositive (27%), and 23 (7%) were undetermined (**Figure S2**). Three seronegative participants were excluded from efficacy analyses, as they were determined post-randomization to be symptomatic at baseline.

Demographics and baseline characteristics were balanced across treatment arms (**Table 1**). For the primary analysis population, mean age was 40.9 years, 45.4% were male, 5.3% identified as Black or African American, and 34.8% identified as Hispanic or Latino. Approximately 71% of seronegative participants had ≥1 risk factor for severe COVID-19 based on the current understanding of these risk factors (**Supplementary Appendix**),^29^ including 62.3% who were overweight (BMI >25 kg/m^2^), 18.6% who had cardiovascular disease or hypertension, and 10.3% who were ≥65 years of age. Similar demographics and characteristics were observed for participants who were seropositive at baseline (**Table S1**).

**Table 1.**
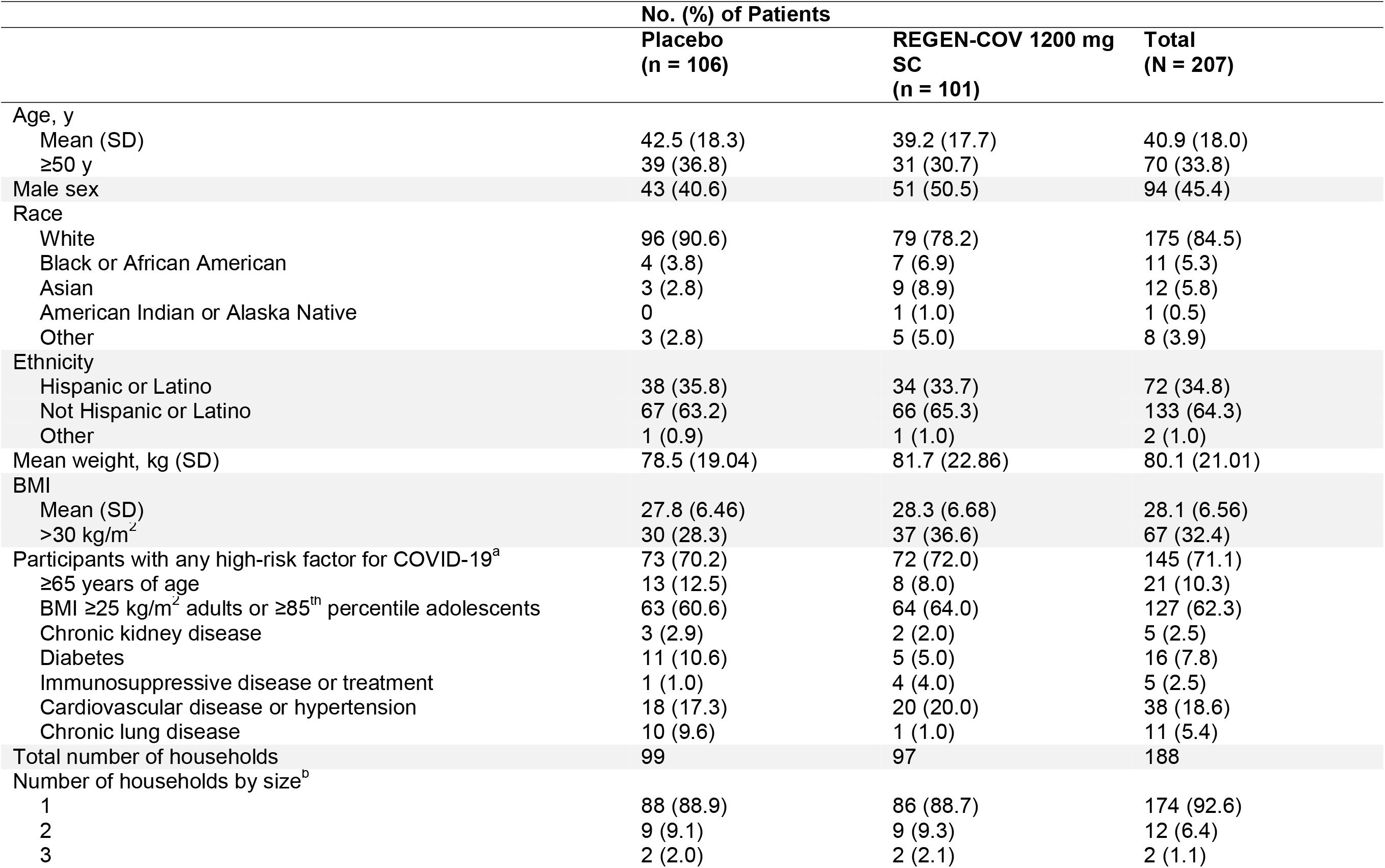
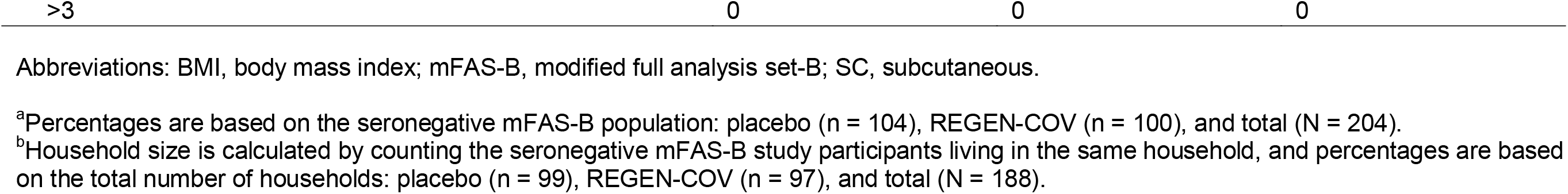
Demographics and Baseline Characteristics (Seronegative).

### Prevention of Symptomatic Infection

In asymptomatic, infected participants who were seronegative, REGEN-COV 1200 mg SC significantly reduced the risk of developing a symptomatic infection (broad-term) compared with placebo (31.5% relative risk reduction; 29/100 [29.0%] vs 44/104 [42.3%]; odds ratio 0.54 [95% CI, 0.30 to 0.97]; *P*=.0380) (**Table 2; Figure 1A**). When symptom onset began 3 days or longer after treatment (day 4 to end of EAP), there was a 76.4% risk reduction in symptomatic infections with REGEN-COV vs placebo (**Table S2**). Findings were similar when utilizing the study-defined strict-term or CDC-defined definitions of COVID-19 signs and symptoms (**Table S3**) and in a post hoc analysis of participants who are more likely to progress to severe COVID-19 based on the current understanding of risk factors for severe disease (**Table S4**).

**Table 2.** Primary and Key Secondary Efficacy Endpoints (Seronegative).^a^.

|  | Placebo<br>(n = 104) | REGEN-COV 1200 mg SC<br>(n = 100) |
| --- | --- | --- |
| Proportion of participants who subsequently develop signs and symptoms (broad-term) within 14 days of a positive RT-qPCR at baseline or during the EAP<sup>b</sup> |  |  |
| No./total no. (%) | 44/104 (42.3) | 29/100 (29.0) |
| Relative risk reduction | - | 31.5% |
| Absolute risk reduction | - | 13.3% |
| Odds ratio (95% CI) | - | 0.54 (0.30–0.97) |
| P value<sup>c</sup> | - | .0380 |
| No. of weeks of symptomatic SARS-CoV-2 infection (broad-term) within 14 days of a positive RT-qPCR at baseline or during the EAP |  |  |
| Total no. of weeks | 170.3 | 89.6 |
| Total per 1000 participants, wk | 1637.4 | 895.7 |
| Reduction<sup>d</sup> | - | 45.3% |
| P value<sup>e</sup> | - | .0273 |
| Mean per-symptomatic participant, wk (SD) | 3.9 (4.5) | 3.1 (4.1) |
| Mean per-participant, wk (SD) | 1.6 (3.5) | 0.9 (2.6) |
| No. of weeks of high viral load (>4 log<sub>10</sub> copies/mL) in NP swab samples during the EAP |  |  |
| Total no. of weeks | 82 | 48 |
| Total duration (wk) per 1000 participants | 811.9 | 489.8 |
| Reduction<sup>d</sup> | - | 39.7% |
| P value<sup>e</sup> | - | .0010 |
| Mean per-participant, wk (SD) | 0.8 (0.8) | 0.5 (0.7) |

Abbreviations: EAP, efficacy assessment period; NP, nasopharyngeal; RT-qPCR, quantitative reverse transcription polymerase chain reaction; SC, subcutaneous.
<sup>a</sup>Three seronegative participants (two in placebo group and one in the REGEN-COV group) were excluded from efficacy analyses, as they were determined post-randomization to be symptomatic at baseline. Key secondary endpoints are presented in order of the hierarchical testing sequence.
<sup>b</sup>Primary endpoint.
<sup>c</sup>Based on logistic regression model adjusted by region (US vs ex-US) and age group (12 to less than 50 years of age vs 50 years of age or older).
<sup>d</sup>Based on the normalized weeks per 1000 participants.
<sup>e</sup>Based on stratified Wilcoxon rank sum test (van Elteren test) with region (US vs ex-US) and age group (12 to less than 50 years of age vs 50 years of age or older) as strata.

**Figure 1.**
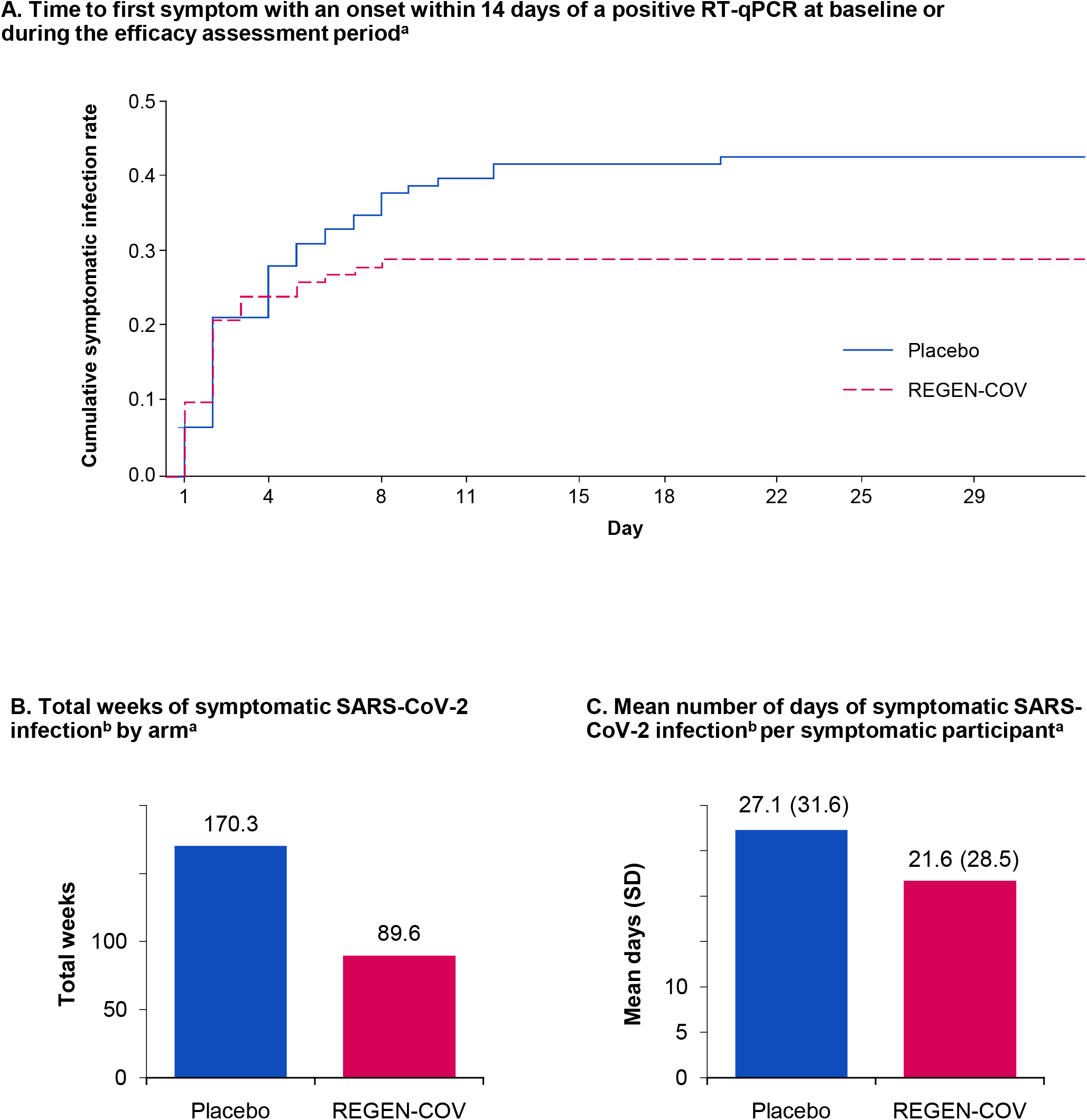
Prevention of Symptomatic SARS-CoV-2 Infection With REGEN-COV. A, Time to first symptom with an onset within 14 days of a positive RT-qPCR at baseline or during the efficacy assessment period.^a^ B, Total weeks of symptomatic SARS-CoV-2 infection^b^ by arm.^a^ C, Mean number of days of symptomatic SARS-CoV-2 infection^b^ per symptomatic participant.^a^ ^a^Seronegative modified full analysis set-B. ^b^Within 14 days of a positive RT-qPCR at baseline or during the efficacy assessment period. Abbreviations: RT-qPCR, reverse transcriptase quantitative polymerase chain reaction; SD, standard deviation.

There was a 45.3% reduction in the aggregated total number of weeks with symptoms (in the overall population) with REGEN-COV versus placebo: 89.6 vs 170.3 weeks or 895.7 vs 1,637.4 weeks per 1000 participants (*P*=.0273; **Table 2; Figure 1B**). This corresponded to a 5.6-day reduction in the mean duration of symptoms per symptomatic participant with REGEN-COV (21.7 days) vs placebo (27.3 days; **Table 2; Figure 1C**).

### Virologic Efficacy

There was a more rapid decline in viral load in REGEN-COV 1200 mg SC–treated participants compared with those treated with placebo, with an adjusted mean difference in viral load of -1.5 log_10_ copies/mL in favor of the antibody combination at Day 8 (**Figure 2A; Figure S3; Table S5**). REGEN-COV treatment also led to a higher proportion of patients with no RT-qPCR–detectable virus at each week during the EAP (**Table S6**).

**Figure 2.**
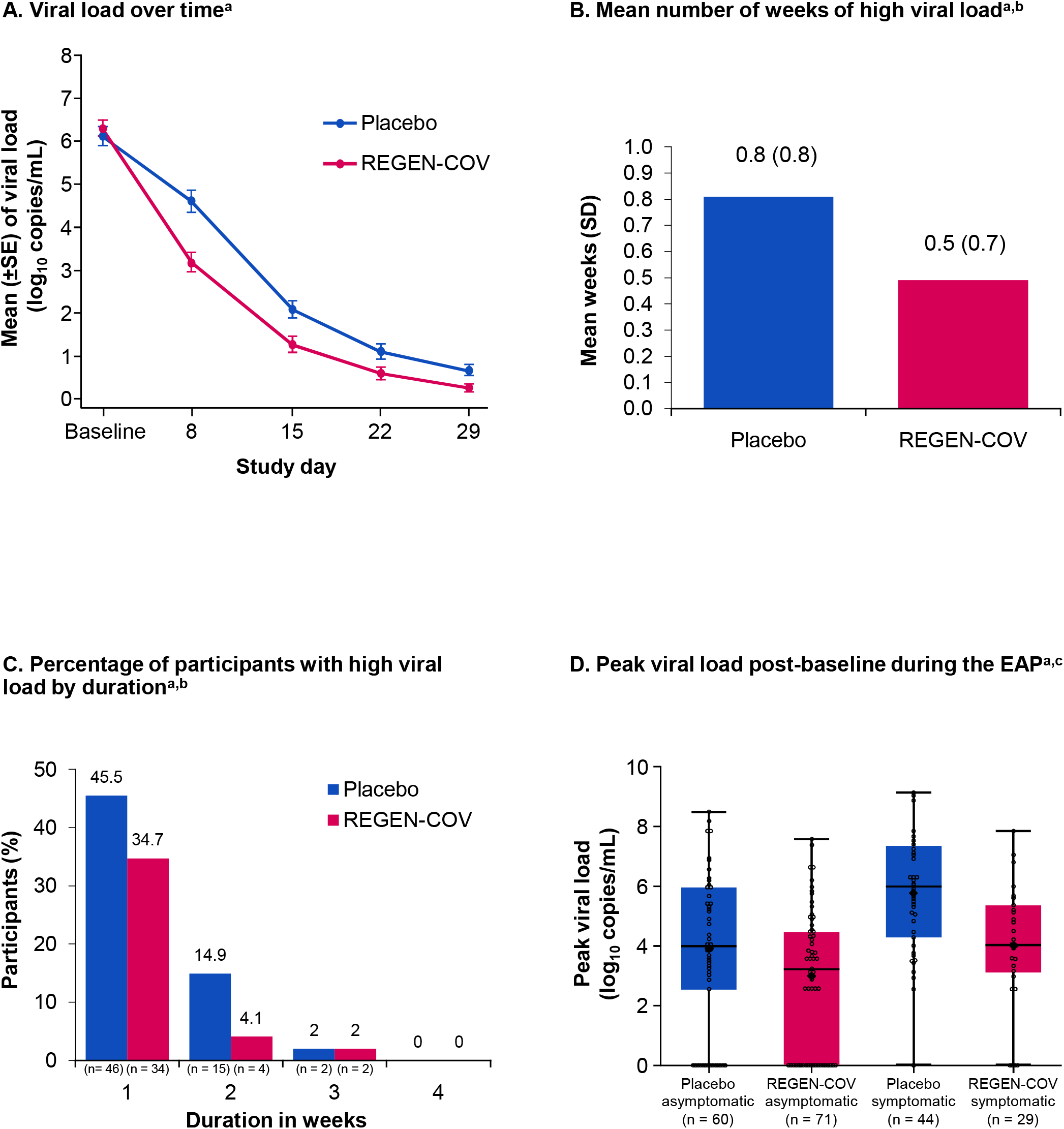
Reduction in SARS-CoV-2 Viral Load With REGEN-COV. A, Viral load over time.^a^ B, Mean number of weeks of high viral load.^a,b^ C, Percentage of participants with high viral load by duration.^a,b^ D, Peak viral load post-baseline during the EAP.^a,c^ ^a^Seronegative modified full analysis set-B. Only non-missing available nasopharyngeal swab viral load data are used for the analysis of viral load endpoints. Only participants with at least one post-baseline viral load data point (in nasopharyngeal swab samples) are included in the analysis. ^b^High viral load was defined as >4 log_10_ copies per milliliter. ^c^Lines in the boxes represent the median; large, bolded dots in the boxes represent the mean; bottom and top of boxes represent quartiles 1 (25th percentile) and 3 (75th percentile), respectively; whiskers represent the maximum and minimum. Abbreviations: EAP, efficacy assessment period; SD, standard deviation; SE, standard error.

The number of weeks of high viral load (>4 log_10_ copies/mL) was significantly reduced with REGEN-COV vs placebo, with a 39.7% reduction in aggregated total number of weeks (in the overall population) (48 vs 82 weeks or 489.8 vs 811.9 weeks per 1000 participants; *P*=.0010; **Table 2**). This corresponded to a 0.3-week reduction per participant in weeks with high viral load from 0.8 weeks with placebo to 0.5 weeks with REGEN-COV (**Table 2; Figure 2B-2C**). Compared with placebo, REGEN-COV reduced peak viral load by approximately 1–2 log_10_ copies/mL both in participants who became symptomatic and in those who remained asymptomatic throughout the EAP (**Figure 2D; Table S5**).

### Prevention of Severe Disease

REGEN-COV 1200 mg SC reduced the proportion of participants who had a COVID-19–related hospitalization or ER visit vs placebo (0/100 vs 6/104, respectively) (**Table S7**). Of the 6 participants in the placebo group, 3 went to the ER, 1 was hospitalized, and 2 went to the ER and were subsequently hospitalized. In contrast, no participants receiving REGEN-COV had ER visits or hospitalizations. Other hospitalization outcomes are provided in **Table S8**.

### Efficacy in Participants According to Baseline Serology Status

Although the primary analysis population focused on participants without evidence of prior infection (seronegative), analyses conducted in all participants combined (seronegative, seropositive, and undetermined) and in the seropositive population analyzed separately showed that REGEN-COV reduced the risk of developing symptomatic infection vs placebo by 35.4% and 33.9%, respectively (**Table S9**). Similarly, in all participants, REGEN-COV reduced the duration of symptoms in those who became symptomatic, reduced the duration of weeks of detectable viral load and high viral load, and reduced peak viral load, with similar numerical trends in the seropositive-only population in most analyses (**Table S10-S13**).

### Safety

REGEN-COV was generally well tolerated. A comparatively lower proportion of participants in the REGEN-COV group experienced TEAEs (33.5%) compared with placebo (48.1%), including both COVID-19-related (25.8% vs. 39.7%, respectively) and non-COVID-19-related (11.0% vs. 16.0%, respectively) events (**Table 3; Table S14**). The most frequent TEAEs and SAEs were COVID-19–related, with a higher proportion of placebo participants with at least one of these events (**Table S14-S15**). Serious TEAEs were reported in 0 participants (0%) in the REGEN-COV group and 4 participants (2.6%) in the placebo group; 1 of the 4 in the placebo group had a non-COVID-19-related serious TEAE (**Table S15**). There were no grade ≥3 injection site reactions or grade ≥3 hypersensitivity reactions in either group (**Table 3**). Injection site reactions (grade 1–2) occurred in 6 participants (4%) in the REGEN-COV group and 1 participant (1%) in the placebo group (**Table S15**). No deaths were reported up to the data cut-off date (March 11, 2021).

**Table 3.** Overview of Treatment-Emergent Adverse Events.

|  | Placebo<br>(n = 156) |  | REGEN-COV 1200 mg SC<br>(n = 155) |  |
| --- | --- | --- | --- | --- |
|  | Overall | Non-COVID-19 | Overall | Non-COVID-19 |
| No. of TEAEs | 109 | 42 | 67 | 26 |
| No. of TEAEs with grade $\geq 3$ | 5 | 2 | 1 | 1 |
| No. of serious TEAEs | 4 | 1 | 0 | 0 |
| No. of AESIs<sup>a</sup> | 0 | 0 | 0 | 0 |
| No. of TEAEs resulting in study drug withdrawal | 0 | 0 | 0 | 0 |
| No. of TEAEs resulting in death | 0 | 0 | 0 | 0 |
| Participants with at least one TEAE, no. (%) | 75 (48.1) | 25 (16.0) | 52 (33.5) | 17 (11.0) |
| Participants with at least one TEAE with grade $\geq 3$, no. (%) | 4 (2.6) | 1 (0.6) | 1 (0.6) | 1 (0.6) |
| Participants with at least one serious TEAE, no. (%) | 4 (2.6) | 1 (0.6) | 0 | 0 |
| Participants with at least one AESI<sup>a</sup>, no. (%) | 0 | 0 | 0 | 0 |
| Participants with at least one TEAE resulting in study drug withdrawal, no. (%) | 0 | 0 | 0 | 0 |
| Participants with at least one TEAE resulting in death, no. (%) | 0 | 0 | 0 | 0 |

Abbreviations: AESI, adverse event of special interest; SC, subcutaneous; TEAE, treatment-emergent adverse event.
<sup>a</sup>Grade $\geq 3$ injection site reactions or hypersensitivity reactions.

### Pharmacokinetics

Following SC administration of a single 1200 mg dose to study participants (in either sentinel or safety groups; for description, see **Supplementary Appendix**), casirivimab and imdevimab were rapidly absorbed, with mean (SD) concentrations in serum one day after dosing of 23.3 (15.0) mg/L and 22.7 (14.8) mg/L; at this timepoint, concentrations were greater than 100 times the IC90 for the delta VOC (**Figure S4; Figure S5**). Both antibodies reached maximal concentrations in serum at a median time of 7.5 days. Casirivimab and imdevimab exhibited linear elimination and had a mean half-life of 30.2 (5.31) days and 26.5 (5.31) days. Mean concentrations in serum 28 days after dosing (C28) were 33.5 (12.3) mg/L and 26.9 (9.12) mg/L for casirivimab and imdevimab. A summary of PK parameters after a single 1200 mg SC dose is shown in **Table S16**. A comparison of antibody concentrations in serum over 28 days following single subcutaneous (this trial) or intravenous doses (two different outpatient trials: NCT04425629 and NCT04666441) of REGEN-COV is shown in **Figure S6**.

## DISCUSSION

REGEN-COV is highly effective for both the treatment and prevention of COVID-19. In symptomatic outpatients with COVID-19 who are at high risk for progression to severe disease, intravenous REGEN-COV 1200 mg has been shown to prevent hospitalization or all-cause death by approximately 70%.^24,25^ Intravenous REGEN-COV 1200 mg also lowered viral load (−0.71 log_10_ copies/mL vs placebo at Day 7) and decreased duration of symptoms (10 vs 14 days placebo) in symptomatic, high-risk outpatients. In part A of the household contact study, we demonstrated that subcutaneous REGEN-COV 1200 mg prevented COVID-19 by approximately 81% when administered to uninfected participants.^28^ In part B of the same household contact study, we now demonstrate that subcutaneous REGEN-COV 1200 mg significantly prevented the progression of asymptomatic SARS-CoV-2 infection to symptomatic COVID-19 by 31.5%. Moreover, REGEN-COV reduced the occurrence of symptomatic disease by more than 75% when symptoms began 3 days or longer after treatment, suggesting a greater impact when treatment is provided earlier in the course of infection and that imminent symptomatic disease is less modifiable. Treatment of asymptomatic, infected participants with REGEN-COV also reduced the duration of high viral load infections by approximately 40%. From a public health perspective, preventive therapy with REGEN-COV to lower SARS-CoV-2 viral load in recently exposed close contacts of infected individuals would be expected to reduce the reservoir for potential further transmission and virus mutation.^30-32^

In the context of a recent high-risk exposure, asymptomatic, infected individuals who have not yet mounted their own humoral immune response (seronegative) constitute the earliest stages of SARS-CoV-2 infection.^33-35^ Consistent with this idea, natural history analyses demonstrated that asymptomatic/PCR-positive/seronegative individuals treated with placebo cleared virus more slowly and were more likely to develop symptomatic disease than asymptomatic/PCR-positive/seropositive individuals treated with placebo (42.3% vs 13.2%, respectively; **Supplementary Results**). These findings are aligned with results from our previously reported outpatient study.^24,25^ Overall, these results indicate that asymptomatic/PCR-positive/seronegative individuals, as observed in this trial, may indeed be earlier in their course of infection and could potentially be key drivers of viral transmission if they remain untreated.^3,5,30^

Although the primary analyses were in seronegative participants, REGEN-COV also provided a benefit in the asymptomatic early infected population, regardless of serostatus. Point-of-care serology tests may therefore have limited utility in guiding decisions in the clinic to prevent COVID-19 in exposed individuals.

The reduction in viral load with subcutaneous REGEN-COV 1200 mg observed in part B of household contact study was comparable to that observed with intravenous REGEN-COV 2400 mg and 1200 mg in a trial in symptomatic outpatients at high risk for progression to severe COVID-19 (**Figure S3**).^24^ Moreover, the reduction in duration of symptoms with subcutaneous REGEN-COV for participants who did become symptomatic in this study (6 days) was similar to that observed in symptomatic outpatients (4 days).^24^ With the majority of participants having risk factors for progression to severe COVID-19 (∼75%), 6 placebo-treated participants had an ER visit or hospitalization while no participants receiving subcutaneous REGEN-COV had these events, consistent with observations showing reduced risk of hospitalization or death in symptomatic outpatients treated with intravenous REGEN-COV.^24^ Overall, these results demonstrate that subcutaneous administration of REGEN-COV 1200 mg has similar antiviral efficacy and clinical benefit as intravenous administration as observed in a study in high-risk outpatients,^24^ supporting the treatment of COVID-19 with either intravenous or subcutaneous administration route, as authorized under the current EUA.^26^

As has been shown consistently in REGEN-COV clinical studies, a larger proportion of participants who received placebo experienced ≥1 TEAE, with the difference attributed to the higher number of COVID-19–related events observed in that group.^24^ Following subcutaneous dosing, concentrations of each antibody in serum were above the predicted neutralization target concentration, based on in vitro and preclinical data,^19-21^ on the first day following dosing and throughout the 28-day efficacy assessment period. A comparison of PK profiles following subcutaneous or intravenous single doses of REGEN-COV (**Figure S6**) supports the potential for use of either route of administration in the treatment of COVID-19, from asymptomatic to symptomatic disease.

### Limitations

This study has several limitations. First, although this study was conducted at multiple sites, all were located in the U.S., Moldova, or Romania, with the majority of sites in the U.S. Second, despite efforts by sites to recruit non-white participants, there were relatively few non-white (self-reported race) participants enrolled; also, few adolescents were enrolled. Third, although the study was adequately powered, the sample size was relatively small due to a study design in which the infection status of asymptomatic participants was not known at inclusion.

## CONCLUSION

REGEN-COV is currently available in the U.S. under EUA for the treatment of mild-to-moderate COVID-19 and for post-exposure prophylaxis in certain individuals exposed to or at high risk of exposure to a SARS-CoV-2–infected individual.^26,27^ Despite widespread vaccination and the authorization of monoclonal antibody combinations, there were approximately 115,000 cases of COVID-19, 10,000 hospitalizations, and 500 deaths in the U.S. every week in August 2021.^36^ Based upon the totality of evidence from phase 3 REGEN-COV clinical studies^24,28^ and previously reported data on REGEN-COV activity against VOC/VOIs,^22^ there is rationale for the use of the antibody combination for treatment as well as for prevention of COVID-19. As a complement to vaccines, widespread utilization of monoclonal antibody combinations that are highly effective against VOCs will be critical to reduce SARS-CoV-2 transmission and COVID-19–related morbidity and mortality.

## Supporting information

Author COI

Author COI

Author COI

Author COI

Author COI

Author COI

Author COI

Author COI

Author COI

Author COI

Author COI

Author COI

Author COI

Author COI

Author COI

Author COI

Author COI

Author COI

Author COI

Author COI

Author COI

Author COI

Author COI

Author COI

Author COI

Author COI

Author COI

Author COI

Author COI

Author COI

Author COI

Author COI

Author COI

Author COI

Author COI

Author COI

Author COI

## Data Availability

Qualified researchers may request access to study documents (including the clinical study report, study protocol with any amendments, blank case report form, statistical analysis plan) that support the methods and findings reported in this manuscript. Individual anonymized participant data will be considered for sharing once the indication has been approved by a regulatory body, if there is legal authority to share the data and there is not a reasonable likelihood of participant re-identification. Submit requests to https://vivli.org/.

## DATA SHARING

A data sharing statement provided by the authors is available with the full text of this article.

## SUPPORTED BY

Supported by Regeneron Pharmaceuticals, Inc. and F. Hoffmann-La Roche Ltd. This trial was conducted jointly with the National Institute of Allergy and Infectious Diseases (NIAID), National Institutes of Health (NIH). The CoVPN is supported by cooperative agreement awards from NIAID, NIH. The work on this study was supported by award numbers UM1AI068619 and UM1AI148684. The content of this manuscript is solely the responsibility of the authors and does not necessarily represent the official views of the NIH.

## ACKNOWLEDGEMENT

We thank the study participants; their families; the investigational site members involved in this trial (listed in the **Supplementary Appendix**); the COVID-19 Phase 3 Prevention Trial Team (listed in the **Supplementary Appendix**); the members of the Data and Safety Monitoring Board; Brian Head, Ph.D., Caryn Trbovic, Ph.D., and S. Balachandra Dass, Ph.D., from Regeneron Pharmaceuticals for assistance with development of the manuscript; and Prime, Knutsford, UK, for formatting and copy editing suggestions.

