## Supplement for "Subcutaneous REGEN-COV Antibody Combination in Early Asymptomatic SARS-CoV-2 Infection: A Randomized Clinical Trial"

**SUPPLEMENTARY APPENDIX**

[Figure S6. Mean (±SD) Concentrations of Casirivimab and Imdevimab in Serum Over Time (Study 2069 [SC*] vs Study 2067 [IV^†^] or 20145 [IV^‡^])	42](#_Toc82779025)

### COVID-19 Phase 3 Prevention Trial Team (REGN 2069/CoVPN 3502)

#### Study Sites and Investigators

**Advanced Pulmonary Research Institute, Loxahatchee, FL:** Neal Warshoff, Liudmila Moreiras

**AGA Clinical Trials, Miami, FL:** Dario Altamirano, Dickson Ellington, Faisal Faikih

**AMR-Knoxville (formerly NOCCR), Knoxville, TN:** William Smith, Richard Gibson, Katie Buckner

**Ardmore Medical Research, Winston Salem, NC:** Robert Rosen, Amy Sapp

**Arizona Liver Health, Tucson, AZ:** Anita Kohli, Vicki McIntyre, Yessica Sachdeva

**Arizona Liver Health, Chandler, AZ:** Yessica Sachdeva, Anita Kohli, Amanda McFarland, Dina Gibson

**Ark Clinical Research, Long Beach, CA:** Kenneth Kim, Jason Ahn, Lisa Neinchel, Nayna Paryani, Amber Mottola, Eva Day, Martha Navarro

**Atella Clinical Research La Palma, CA:** Rafaelito Victoria, Xanthe Victoria, Rene Uong

**Atrium Health, Charlotte, NC:** Mindy Sampson, Christopher Polk, Michael Leonard, Lewis McCurdy, Leigh A. Medaris, Zainab Shahid, Lisa Davidson

**Avera McKennan Hospital and University Health Center, Sioux Falls, SD:** Jawad Nazir, John Lee, Amy Elliott, Swami Sathyanaryan, Mansi Oberoi, Muhammad (Danial) Siddiqui, Muhammad Arsad, Kara Bruning

**Adolescent & Young Adult Research at Core – NIAID CoVPN, Chicago, IL:** Sybil Hosek, Temitope Oyedele, Vanessa Sarda, Monica Mercon

**Beth Israel Deaconess Medical Center, Boston, MA:** Kathryn Stephenson, Dan Barouch, Boris Juelg, Chen Sabrina Tan, Rebecca Zash, Ai-ris Collier, Jessica Ansel, Kate Jaegle

**Bio-Medical Research, LLC, Miami, FL:** Lilia Roque-Guerrero, Ana Gomez Ramirez, Javier Capote, Gisel Paz

**Boston Medical Center Ped. HIV Program NICHD – NIAID CoVPN, Boston, MA:** Michael Paasche-Orlow, Julien Dedier

**California Medical Research Associates, Northridge, CA:** Sanjay Vadgama, Ramachandra Patak

**Cardiology Care Clinics, Eatonton, GA:** Nicolas Chronos, Cary Hefty

**Carolina Institute for Clinical Research, Fayetteville, NC:** Judith Borger, Ifeanyi Momodu, Lindsey Carswell, Benjamin King, Ryan Starr, Scott Syndergaard

**Carolina Medical Research, Clinton, SC:** Nancy Patel, Ravikumar Patel, Ryan Sattar

**Catalina Research Institute, Montclair, CA:** Rizwana Mohseni, Jeffrey Unger, Sheila De Jesus-Maranan, Cecilia Casaclang

**Centex Studies, Lake Charles, LA:** Michael Seep, Celeste Brown, Joshua Whatley

**Chicago Clinical Research Institute, Chicago, IL:** Dennis Levinson, Saad Alvi, Norman James, Azazuddin Ahmed

**Clinical Research of Central Florida, Winter Haven, FL:** Robinson Koilpillai, Stephanie Cassady, Jennifer Cox, Eduardo Torres

**Clinical Trial of Florida LLC, Miami, FL:** James Krainson, Mark J. Rosenthal

**Crossroads Clinical Research, Corpus Christi, TX:** Michael Winnie, Jerry Plemons, Omesh Verma, Richard Leggett

**DM Clinical Research/BFHC Research, San Antonio, TX:** Ramon Reyes, Keith Beck, Brian Poliquin

**DM Clinical Research/LinQ Research, LLC, Pearland, TX:** Murtaza Mussaji, Jignesh Shah

**East Coast Institute for Research, Jacksonville, FL:** David Sutton, Edward Pereira, Rodel Gloria, Stacey Kelly, Amy Dennis-Saltz, Mae Sheikh-Ali, Elias Saikali, James Magee, Rebecca Goldfaden

**Epic Medical Research, Red Oak, TX:** Haresh Boghara, Sunny Patel, Bari Eichelbaum

**Excel Clinical Research, Las Vegas, NV:** Duane Anderson, Sean Su, Alexander Akhavan, Diana Kirby, Joy Venglik

**Fenway Health – NIAID CoVPN, Boston, MA:** Kenneth Mayer, Taimur Khan, Marcy Gelman

**Florida Pulmonary Research Institute, LLC, Winter Park, FL****:** Faisal A. Fakih, Faisal M. Fakih, Daniel Layish, Fernando Alvarado, Jose Diaz

**Fomat Medical Research, Oxnard, CA:** Augusto Focil, Griselda Rosas, Stevan Correa, Michael Bogseth

**Future Innovative Treatments, LLC, Colorado Springs, CO:** Bhaktasharan Patel, Gary Tarshis, Katrina Grablin

**Geisinger Medical Center, Danville, PA:** Paul Simonelli, Stanley Martin, Alvin Sharma, Anna Chen, Pragya Dhaubhadel, Shaeesta Khan, Sreelatha Naik, Sudheer Penupolu, Thulashie Sivarajah, Tae-Sung Kwon, Lakshmi Saladi

**Geisinger Wyoming Valley, Wilkes-Barre, PA:** Paul Simonelli, Stanley Martin, Alvin Sharma, Anna Chen, Pragya Dhaubhadel, Shaeesta Khan, Sreelatha Naik, Sudheer Penupolu, Thulashie Sivarajah, Tae-Sung Kwon, Lakshmi Saladi

**Harlem Hospital Center – New York City Health and Hospitals Corporation, Harlem, NY:** Farbod Raiszadeh, Sharon Mannheimer, Khaing Myint, Hussein Assallum, Lovelyamma Varghese, Akari Kyawa

**Harlem Prevention Center – NIAID CoVPN, Harlem, NY:** Ellen Morrison, Sharon Mannheimer, Julie Franks, Jun Avelino Loquere, Orlando Rosario, Andrea Low, Joan Villacrucis

**HD Research Group, Houston, TX:** Alan Skolnick, Harold Minkowitz, David Leiman, Todd Price, Anatoli Krasko, Idisoro Wiener

**Healthcare Research Network, Hazelwood, MO:** Larry Reed, Oscar Lin

**Henry Ford Health System, Detroit, MI:** Mayur Ramesh, George Alangaden

**Holy Name Medical Center, Teaneck, NJ:** Suraj Saggar, Thomas Birch, Benjamin De La Rosa, Karyna Neyra, Erina Kunwar

**IACT Health, Columbus, GA:** Jeffrey Kingsley, April Pixler, Veronica McBride

**Icahn School of Medicine at Mount Sinai, New York, NY:** Judith Aberg, Michelle Cespedes, Alexandra Abrams-Downey, Erna Kojic, Luz Lugo, Sean Liu, Nadim Salomon, David Perlman, Deena Altman, Farah Rahman, Georgina Osorio, Joseph Mathew, Sanjana Koshy, Dana Mazo, Francesca Cossarini, Sondra Middleton, Alina Jen, Erika Maria Reategui Schwarz

**Innovative Research of West Florida, Clearwater, FL:** Miguel Trevino, Benjamin DeVries

**Lincoln Medical Center – New York City Health and Hospitals Corporation, Bronx, NY:** Vidya Menon, Moiz Kasubhai, Usha Venugopal, Anjana Pillai, Franscene Oulds

**M3 Wake Research, Raleigh, NC:** Matthew Hong, Wayne Harper, Lynn Eckert, Douglas Wadeson, Lisa Cohen

**Maryland School of Medicine, Baltimore, MD:** Joel Chua, Shyam Kottilil, Jennifer Husson, John Baddley, R. Gentry Wilkerson, Shivakumar Narayanan, Uzoamaka Eke, Myint Noe, Melanie Malave Sanchez

**Massachusetts General Hospital – ID Clinical Research Unit – NIAID CoVPN, Boston, MA:** Arthur Kim, Greg Robbins, Mark Siedner, Rajesh Gandhi, Kristen Hysell, Jacob Lazarus, Lael Yonker

**McGovern Medical School at The University of Texas Health Science Center, Houston, TX:** Roberto Arduino, Karen J. Vigil

**Medical Research of Westchester, Miami, FL:** Richard Perez-Perez, Carlos J. Bello, Esperanza Arce-Nunez, Jorge Acosta, Julio L. Arronte

**Medical University of South Carolina, Charleston, SC:** Eric Meissner, Patrick Flume, Andrew Goodwin, Deeksha Jandhyala, Nandita Nadig

**MedPharmics, Metairie, LA:** Robert Jeanfreau, Susan Jeanfreau, Susan Tortorich, Shiva Akula

**MedPharmics, Gulfport, MS:** Paul Matherne, Donald Gaddy, Magdy Mikhail

**Mercury Clinical Research, Houston, TX:** Rajasekaran Annamalai, Huy Nguyen, Nizar Nayani, Mahalakshmi Ramchandra

**META Medical Research Institute, Dayton, OH:** Priyesh Mehta, Jacqueline Horne, Grace Hassan

**Midland Florida Clinical Research Center, Deland, FL:** Godson Oguchi, Judepatricks Onyema

**Midway Immunology and Research Center, Fort Pierce, FL:** Moti Ramgopal, Brenda Jacobs, Lisa Cason, Angela Trodglen

**National Institute of Infectious Diseases, Bucharest, Romania:** Adrian Streinu, Daniela Manolache, Anca Streinu-Cercel, Oana Sandulescu, Ana Blanaru, Monica Stoica, Ana Maria Andone, Daniela Dospinoiu, Silviu Serban, Loredana Patru, Christina Buhuara, Ramona Dorobantu, Magdalena Motoi, Ioana Daramus, George Bihoi, Alexandra Ghita, Victor Miron, Gylda Spataru

**New Jersey Medical School Clinical Research Center – NIAID CoVPN, Newark, NJ:** Amesika Nyaku, Shobha Swaminathan

**Next Level Urgent Care, Houston, TX:** Terence Chang, Robbyn Traylor, Lenee Gordon, John McDivitt, Lizette Castro

**Northern California Research, Sacramento, CA:** Douglas Young, Gary Carson

**New York University Langone Vaccine Center – NIAID CoVPN, Manhattan, NY:** Angelica Kottkamp, Mark J. Mulligan, Anna Bershteyn, Vanessa Raabe, Tamia Davis, Mary Olson

**Ohio State University AIDS Clinical Trials Unit – NIAID CoVPN, Columbus, OH:** Seuli Brill, Carlos Malvestutto, Susan Koletar, Taru Saigal, Mahdee Sobhanie, Vignesh Doraiswamy, Mahrous Abo Hassan, Jeremy Young

**Orlando Immunology Center, Orlando, FL:** Edwin DeJesus, Charlotte-Paige Rolle, Federico Hinestrosa, Dan Cruz, Terry Wilder, Jeffrey Garrett, Stephanie Skipper

**Paradigm Clinical Research Institute, Torrance, CA:** Ramprasad Dandillaya, Kartik Ananth

**Penn Prevention – NIAID CoVPN, Philadelphia, PA:** Ian Frank, Helen Koenig, Eileen Donaghy, Debora Dunbar

**PMG Research of McFarland Clinic, Ames, IA:** Jennifer Killion, Rupal Amin, Shauna Basener, Timothy Lowry

**PMG Research of Wilmington, Wilmington, NC:** Kevin Cannon, Mesha Chadwick

**Qway, Hialeah, FL:** Oscar Galvez, Fausto Castillo

**Regional One Health, Memphis, TN:** John Jefferies, Sandy Arnold, Amber Thacker

**Remington-Davis, Columbus, OH:** Edward Cordasco, Brian Zeno, Heather Holmes, Heather Lee

**Republican Clinical Hospital, Chisinau, Moldova:** Natalia Gaibu, Victor Cojocaru, Aristia Seremet, Sergiu Iacob, Rodica Usatii, Nelea Ghicavii, Angela Coltuclu, Oxana Bujor

**Rhode Island Hospital, Providence, RI:** Eleftherios Mylonakis, Dimitrios Farmakiotis, Karen Tashima, Natasha Ryback

**Ruane Clinical Research Group, Los Angeles, CA:** Peter Ruane, Peter Wolfe, Kenny Trinidad

**Rush University Medical Center, Chicago, IL:** James Moy, Raj Shah, Bandi Sindhura, Beverly Sha

**San Francisco Research Institute, San Francisco, CA:** Mark Savant, Francis Hsiao, Edna Yee

**Sarasota Memorial Hospital, Sarasota, FL:** Manuel Gordillo, Rishi Bhattacharyya, Sudha Tallapragada, Annette Artau, Julie Larkin, Roberto Mercado, Michael Milam, Natan Kraitman, Michael Lowry, Sarah Temple, Lenka Offner, Rabih Loutfi, Kirk Voelker, Marshall Frank, Ashley Grant

**SignatureCare Emergency Center – TC Jester, Houston, TX:** Alan Skolnick, Harold Minkowitz, David Leiman, Todd Price, Anatoli Krasko

**St. Hope Foundation, Bellaire, TX:** James Sims III, Manuel Vasquez, Kenneth Degazon, Katherine Asuncion

**Stanford University, Palo Alto, CA:** Jason Andrews, Aruna Subramanian, Upinder Singh, Yvonne Maldonado, Chaitan Khosla

**Tandem Clinical Research, Maitland, FL:** Esteban Olivera, Mayra Abreu

**Tandem Clinical Research, Marrero, LA:** Adil Fatakia, Marissa Miller, Kristen Clinton, Gary Reiss

**The Hope Clinic of Emory University – NIAID CoVPN, Decatur, GA:** Srilatha Edupuganti, Nadine Rouphael, Colleen Kelley, Varun Phadke, Cassie Grimsley Ackerley, Matthew Collins

**The Lundquist Institute, Torrance, CA:** Loren Miller, Timothy Hatlen

**The Ponce de Leon Center Clinical Research Site – NIAID CoVPN, Atlanta, GA:** Michael Chung, Colleen Kelley, Valeria Cantos Lucio, Carlos del Rio, Jeffrey Lennox, Sheetal Kandiah, Caitlin Moran, Anandi Sheth, Paulina Rebolledo, Nithin Gopalsamy, Divya Bhamidipati

**Triple O Research Institute PA, West Palm Beach, FL:** Olayemi Osiyemi, Jose A. Menajovsky-Chaves, Christina Campbell

**Tufts Medical Center, Boston, MA:** Andrew Strand, Andreas Klein, Debra Poutsiaka, Roberto Viau Colindres, Brian Chow, Cheleste Thorpe, Mary Hopkins, Jenn Chow, Rakhi Kohli, Jose Caro, Jeffrey Griffiths, Helen Boucher, Whitney Perry, Laura Kogelman, Yoav Golan, Tine Vindenes, Carlos Mendoza, Saba Mostafavi, Christhian Alejandro Cano Guerra, Paula Dabenigno, Bipin Malla

**Tulane University School of Medicine, New Orleans, LA:** Dahlene Fusco, Arnaud Drouin, Joshua Denson, Jerry Zifodya, Christine Bojanowski, Monika Dietrich, Stacy Drury

**University of Illinois at Chicago Project WISH – NIAID CoVPN, Chicago, IL:** Jesica Herrick, Richard Novak, Mahesh Patel

**Universal Medical and Research Center, LLC, Miami, FL:** Gerard Acloque, Agustin Martinez

**University at Buffalo, State University of New York, Buffalo, NY:** Sanjay Sethi, Brian Clemency, Rajesh Kunadharaju

**University of Arizona, Tucson, AZ:** Sairam Parthasarathy, Franz Rischard

**University of California Davis, Sacramento, CA:** Stuart Cohen, George Thompson, Hien Nguyen, Scott Crabtree

**University of Cincinnati, Cincinnati, OH:** Carl Fichtenbaum, Moises Huaman, Jaime Robertson

**University of Colorado School of Medicine, Aurora, CO:** Eric Simoes, Thomas Campbell, Poornima Ramanan, Hillary Dunlevy, Esther Benamu, Amiran Baduashvili, Martin Krsak, Steven Johnson, Lakshmi Chauhan, Erica Fredregil, Samantha Economos

**University of Miami – Miller School of Medicine, Miami, FL:** Gary Kleiner, Lilian Abbo, Bhavarth Shukla, Jennifer Gebbia, Maria Rodriguez

**University of Minnesota, Minneapolis, MN:** Anne-Marie Leuck, Mahsa Abassi, Matthew Pullen

**University of Mississippi, Jackson, MS:** Jose Lucar Lloveras, Leandro Mena, Luis Shimose Ciudad

**University of North Carolina, Chapel Hill, NC:** Jessica Lin, David Wohl, Christopher Hurt, William Fischer II, Kathleen Tompkins

**University of South Florida, Tampa, FL:** Kami Kim, Seetha Lakshmi, Charurut Somboonwit, Jason Wilson, Asa Oxner, Tiffany Vasey, Lucy Guerra

**University of Virginia, Charlottesville, VA:** William Petri, Katie Dykstra, Marianne Morrissey, Lejla Cesko, Jae Shin, Cirle Warren, Jennifer Sasson, Chelsea Marie, Debbie-Ann Shirley, Rebecca Carpenter, Gregory Madden, Danielle Donigan, Michelle Sutton, Cynthia Edwards, Elizabeth Brooks, Rebecca Wade, Samantha Simmons, Jennifer Pinnata

**University of Washington Medical Center, Seattle, WA:** Ruanne Barnabas, Shelly Karuna, Ann C. Collier, Julie McElrath, Janine Maenza, Adrienne Shapiro, Helen Stankiewicz-Karita, Helen Chu, Chandler Church

**University of Wisconsin, Madison, WI:** William Hartman, Joseph Connor, Robert Striker

**University of Texas Health Science Center, Tyler, TX:** Julie Philley, Megan Devine, Richard Yates, Steven Hickerson

**Vanderbilt Vaccine Clinical Research Site – NIAID CoVPN, Nashville, TN:** Spyros Kalams, Greg Wilson

**Virginia Commonwealth University School of Medicine, Richmond, VA:** Michael Donnenberg, Marjolein de Wit

**VitaLink Research, Gaffney, SC:** David Erb, Luis DeLaCruz, Supinder Channa

**Whitman-Walker Health – NIAID CoVPN, Washington, DC:** Sarah Henn, Megan Coleman, Lynsay MacLaren, Deborah Goldstein, Alice Eggleston, Carrington Koebele

**WR-ClinSearch, LLC, Chattanooga, TN:** Mark McKenzie, Teresa Deese

**WR-Mount Vernon Clinical Research, LLC, Sandy Springs, GA:** Benjamin Thomas, Laura Tsakiris, Stephen Blank, Ronald Mirenda

**Xera Med Research, Boca Raton, FL:** Anna Martin, Gargi Gharat, Candace Kokaram, Ket Wray, Clement Partap, Ulyana Arzamasova, Kristina Louissaint, Maria Fernandez

**Xera Med Research, Miami, FL:** Anna Martin, Ket Wray, Kristina Louissaint, Maria Fernandez, Gargi Gharat

#### Regeneron Study Team

Achint Chani, Adebiyi Adepoju, Adnan Mahmood, Aisha Mortagy, Ajla Dupljak, Alina Baum, Alison Brown, Amy Froment, Andrea Hooper, Andrea Margiotta, Andrew Bombardier, Anita Islam, Anne Smith, Arvinder Dhillon, Audra McMillian, Aurora Breazna, Ayesha Aslam, Barabara Carpentino, Bari Kowal, Barry Siliverstein, Benjamin Horel, Bo Zhu, Bret Musser, Brian Bush, Brian Head, Brian Snow, Bryan Zhu, Camille Debray, Careta Phillips, Carmella Simiele, Carol Lee, Carolyn Nienstedt, Caryn Trbovic, Casey (Kuo-Chen) Chan, Catherine Elliott, Chad Fish, Charlie Ni, Christa Polidori, Christine Enciso, Christopher Caira, Christopher Powell, Christos A. Kyratsous, Cliff Baum, Colin McDonald, Cynthia Leigh, Cynthia Pan, Dana Wolken, Danielle Manganello, David Liu, David Stein, David M. Weinreich, Dawlat Hassan, Daya Gulabani, Deborah Fix, Deborah Leonard, Deepshree Sarda, Denise Bonhomme, Denise Kennedy, Devin Darcy, Dhanalakshmi Barron, Diana Hughes, Diana Rofail, Dipinder Kaur, Divya Ramesh, Dona Bianco, Donna Cohen, Eduardo Forleo-Neto, Edward Jean-Baptiste, Ehsan Bukhari, Eileen Doyle, Elizabeth Bucknam, Emily Labriola-Tomkins, Emily Nanna, Esther Huffman O'Keefe, Evelyn Gasparino, Evonne Fung, Flonza Isa, Fung-Yee To, Gary Herman, George D. Yancopoulos, Georgia Bellingham, Giane Sumner, Grainne Moggan, Grainne Power, Haixia Zeng, Hazel Mariveles, Heath Gonzalez, Helen Kang, Hibo Noor, Ian Minns, Ingeborg Heirman, Izabella Peszek, James Donohue, Jamie Rusconi, Janice Austin, Janie Parrino, Jeannie Yo, Jenna McDonnell, Jennifer D. Hamilton, Jessica Boarder, Jianguo Wei, Jingchun Yu, Joanne Malia, Joanne Tucciarone, Jodie Tyler-Gale, John D. Davis, John Strein, Jonathan Cohen, Jonathan Meyer, Jordan Ursino, Joseph Im, Joseph Tramaglini, Joseph Wolken, Kaitlyn Potter, Kaitlyn Scacalossi, Kamala Naidu, Karen Browning, Karen Rutkowski, Karen Yau, Katherine Woloshin, Kelly Lewis-Amezcua, Kenneth Turner, Kimberly Dornheim, Kit Chiu, Kosalai Mohan, Kristina McGuire, Kristy Macci, Kurt Ringleben, Kusha Mohammadi, Kyle Foster, Latora Knighton, Leah Lipsich, Lindsay Darling, Lisa Boersma, Lisa Cowen, Lisa Hersh, Lisa Jackson, Lisa Purcell, Lisa Sherpinsky, Livia Lai, Lori Faria, Lori Geissler, Louise Boppert, Lyra Fiske, Marc Dickens, Marco Mancini, Maria C. Leigh, Meagan O'Brien, Michael Batchelder, Michael Klinger, Michael Partridge, Michel Tarabocchia, Michelle Wong, Mivianisse Rodriguez, Moetaz Albizem, Muriel O'Byrne, Ned Braunstein, Neena Sarkar, Neil Stahl, Nicole Deitz, Nicole Memblatt, Nirav Shah, Nitin Kumar, Olga Herrera, Oluchi Adedoyin, Ori Yellin, Pamela Snodgrass, Patrick Floody, Paul D'Ambrosio, Paul (Xiaobang) Gao, Peijie Hou, Philippa Hearld, Qin Li, Rachel Kitchenoff, Rakiyya Ali, Ramya Iyer, Ravikanth Chava, Rinol Alaj, Rita Pedraza, Robert Hamlin, Romana Hosain, Ruchin Gorawala, Ryan White, Ryan Yu, Rylee Fogarty, S. Balachandra Dass, Sagarika Bollini, Samit Ganguly, Sandra DeCicco, Sanket Patel, Sarah Cassimaty, Selin Somersan-Karakaya, Shane McCarthy, Sharon Henkel, Shazia Ali, Shelley Geila Shapiro, Somang Kim, Soraya Nossoughi, Stephanie Bisulco, Steven Elkin, Steven Long, Sumathi Sivapalasingam, Susan Irvin, Susan Wilt, Tami Min, Tatiana Constant, Theresa Devins, Thomas DiCioccio, Thomas Norton, Travis Bernardo, Tzu-Chien Chuang, Victor (Jianguo) Wei, Vinh Nuce, Vishnu Battini, Wilson Caldwell, Xiaobang Gao, Xin Chen, Yanmei Tian, Yasmin Khan, Yuming Zhao, Yunji Kim

#### NIAID/CoVPN Team

Bonnie Dye (CoVPN), Christopher B. Hurt (CoVPN), Dale R. Burwen (NIAID), Dan H. Barouch (CoVPN), David Burns (NIAID), Elizabeth Brown (CoVPN), Katharine J. Bar (CoVPN), Mary Marovich (NIAID), Meredith Clement (CoVPN), Myron S. Cohen (CoVPN), Nirupama Sista (CoVPN), Ruanne V. Barnabas (CoVPN), Sheryl Zwerski (NIAID)

### Supplementary Methods

#### Trial Oversight

Regeneron Pharmaceuticals, Inc. (Regeneron) designed the trial in collaboration with CoVPN and NIAID, and gathered the data with the trial investigators. Regeneron analyzed the data. The investigators, site personnel, CoVPN, NIAID, and Regeneron were blinded to treatment-group assignments. A Data and Safety Monitoring Board convened by the National Institutes of Health evaluated safety data to make recommendations for trial modification and/or termination.

The trial was conducted in accordance with the principles of the Declaration of Helsinki, Good Clinical Practice /International Conference on Harmonisation-E-9 guidelines, and all applicable regulatory standards. The central or local institutional review board or ethics committee at each study center oversaw trial conduct and documentation. All participants provided written informed consent before participating in the trial.

#### Trial Design – Staggered Enrollment

To maximize efficient enrollment while optimizing the safety of eligible participants, enrollment proceeded in a staggered manner by subset as follows.

 **Sentinel group (subset 1; first 30 adult participants [age ≥18 years])** had samples collected in the efficacy assessment period (EAP) and follow-up periods for clinical laboratory tests (hematology, blood chemistry, urinalysis), drug concentration measurement (dense pharmacokinetic [PK] sampling), and immunogenicity analysis (anti-drug antibodies). These participants were enrolled into Part A or Part B as appropriate.
 **Safety group (subset 2; 31st to 400th adult participant)** had samples collected in the EAP and follow-up periods for clinical laboratory tests (hematology, blood chemistry, urinalysis), drug concentration measurement (sparse PK sampling), and immunogenicity analysis (anti-drug antibodies). These participants were enrolled into Part A or Part B as appropriate.
 **All other adult and adolescent participants (subset 3; 401st to last adult or adolescent participant [age ≥12 years])** had samples collected at baseline and at the end of the EAP for clinical laboratory tests (hematology, blood chemistry, urinalysis) and anti-drug antibodies. These participants were enrolled into Part A or Part B as appropriate.

#### Stratification

Assignment of treatment group (1200 mg REGEN-COV or placebo) was stratified by the following prior to randomization:

1. Test results (positive, negative, or undetermined) of a local diagnostic assay for SARS-CoV-2 (eg, molecular assay such as RT-PCR assay for SARS-CoV-2 or a SARS-CoV-2 antigen test) from appropriate samples (eg, nasopharyngeal [NP], oropharyngeal, nasal, or saliva)
2. Age:
   - ≥12 and <18 years
   - ≥18 and <50 years
   - ≥50 years

#### Inclusion and Exclusion Criteria

**Inclusion criteria:**

A participant must meet the following criteria to be eligible for inclusion in the study:

1. Adult participants ≥18 years of age (irrespective of weight) at the signing of informed consent, or adolescent participants ≥12 to <18 years of age, or pediatric participants <12 years of age at the signing of the assent (parent/guardian signs the informed consent)
2. Asymptomatic household contact with exposure to an individual with a diagnosis of SARS-CoV-2 infection (index case). To be included in the study, participants must be randomized within 96 hours of collection of the index cases’ positive SARS-CoV-2 diagnostic test sample
3. Anticipates living in the same household with the index case until study day 29
4. Judged by the investigator to be in good health based on medical history and physical examination at screening/baseline, including participants who are healthy or have a chronic, stable medical condition
5. Is willing and able to comply with study visits and study-related procedures/assessments
6. Provides informed consent signed by study participant or legally acceptable representative

**Exclusion criteria:**

A participant who meets any of the following criteria will be excluded from the study:

1. Participant-reported history of prior positive SARS-CoV-2 RT-PCR test or positive SARS CoV-2 serology test at any time before the screening
2. Participant has lived with individuals who have had previous SARS-CoV-2 infection or currently lives with individuals who have SARS-CoV-2 infection, with the exception of the index case(s), the first individual(s) known to be infected in the household
3. Active respiratory or non-respiratory symptoms consistent with COVID-19
4. History of respiratory illness with sign/symptoms of SARS-CoV-2 infection, in the opinion of the investigator, within the prior 6 months to screening
5. Nursing home resident
6. Any physical examination findings, and/or history of any illness, concomitant medications, or recent live vaccines that, in the opinion of the study investigator, might confound the results of the study or pose an additional risk to the participant by their participation in the study
7. Current hospitalization or was hospitalized (ie, >24 hours) for any reason within 30 days of the screening visit
8. History of significant multiple and/or severe allergies (eg, latex gloves), or has had an anaphylactic reaction to prescription or non-prescription drugs or food. This is to avoid possible confounding of the safety analysis and not due to any presumed increased risk of these individuals to a reaction to the investigational product
9. Treatment with another investigational agent in the last 30 days or within 5 half-lives of the investigational drug, whichever is longer, prior to the screening visit
10. Received an investigational or approved SARS-CoV-2 vaccine
11. Received investigational or approved passive antibodies for SARS-CoV-2 infection prophylaxis (eg, convalescent plasma or sera, monoclonal antibodies, hyperimmune globulin)
12. Use of hydroxychloroquine/chloroquine for prophylaxis/treatment of SARS-CoV-2 or anti-SARS-viral agents,^a^ eg, remdesivir, within 60 days of screening

^a^Hydroxychloroquine/chloroquine for other uses, ego, for use in autoimmune diseases is allowed

13. Member of the clinical site study team and/or immediate family
14. Exclusion criterion #14 excluding sexually active men who are unwilling to use medically acceptable birth control (vasectomy with medical assessment of surgical success OR consistent use of a condom) during the study drug follow-up period and for 8 months after single dose of study drug was removed since enrollment was expanded to include all women in protocol amendment 4.
15. Exclusion criterion #15 excluding pregnant or breastfeeding women was removed since enrollment was expanded to all women in protocol amendment 4.
16. Exclusion criterion #16 excluding women of childbearing potential (WOCBP)^b^ and girls at or beyond menarche (≥12 to <18 years of age) who were unwilling to practice highly effective contraception prior to the initial dose/start of the first treatment, during the study, and for at least 8 months after the last dose was removed since enrollment was expanded to all women in protocol amendment 4.

^b^WOCBP are defined as women who are fertile following menarche until becoming postmenopausal, unless permanently sterile. Permanent sterilization methods include hysterectomy, bilateral salpingectomy, and bilateral oophorectomy.

#### Risk Factors for Severe COVID-19

The following are risk factors for severe COVID-19 (<https://www.cdc.gov/coronavirus/2019-ncov/need-extra-precautions/people-with-medical-conditions.html>. Accessed August 2021)

CDC, August 2021):

- ≥65 years of age
- Body mass index ≥25 kg/m^2^ for adults or ≥85^th^ percentile for adolescents
- Chronic kidney disease
- Diabetes
- Immunosuppressive disease or treatment (ie, immunocompromised state)
- Cardiovascular disease or hypertension
- Chronic lung disease

#### COVID-19 Signs and Symptoms – Broad-Term, Strict-Term, and CDC Definitions

The assessment of association of symptomatic disease with SARS-CoV-2 infection was based on the broad-term, strict-term, and Centers for Disease Control and Prevention (CDC) COVID-19 definitions described below. In order to ensure all symptomatic SARS-CoV-2 infections were captured, the broad-term definition was used for the primary analysis; strict-term and CDC definitions were used in other analyses.

**Broad-Term**

Fever ≥38°C

The signs and symptoms below:

1. Feverish

2. Sore throat

3. Cough

4. Shortness of breath/difficulty breathing *(nasal flaring*^a^*)*

5. Chills

6. Nausea

7. Vomiting

8. Diarrhea

9. Headache

10. Red or watery eyes *(conjunctivitis)*

11. Body aches such as muscle pain or joint pain *(myalgia, arthralgia)*

12. Loss of taste/smell

13. Fatigue *(fatigue or general malaise or lethargy* ^a^*)*

14. Loss of appetite or poor eating/feeding

15. Confusion

16. Dizziness

17. Pressure/tightness in chest

18. Chest pain

19. Stomach ache (abdominal pain^a^)

20. Rash

21. Sneezing

22. Runny nose

23. Sputum/phlegm

Other

*^a^Signs and symptoms observed in pediatric participants*

**Strict-Term**

Fever (≥38°C) PLUS ≥1 respiratory symptom (sore throat, cough, shortness of breath)

*OR*

2 respiratory symptoms (sore throat, cough, shortness of breath)

*OR*

1 respiratory symptom (sore throat, cough, shortness of breath) PLUS ≥2 non-respiratory symptoms (chills, nausea, vomiting, diarrhea, headache, conjunctivitis, myalgia, arthralgia, loss of taste or smell, fatigue or general malaise)

**CDC Definition**

*https://www.cdc.gov/coronavirus/2019-ncov/symptoms-testing/symptoms.html*

At least 2 of the following symptoms: fever (measured or subjective), chills, rigors, myalgia, headache, sore throat, nausea or vomiting, diarrhea, fatigue, congestion or runny nose

OR

Any 1 of the following symptoms: cough, shortness of breath, difficulty breathing, new olfactory disorder, new taste disorder

OR

Severe respiratory illness with at least 1 of the following, clinical or radiographic evidence of pneumonia, acute respiratory distress syndrome.

#### Pharmacokinetic Analysis Methods

**Bioanalytical Methods**

The concentrations of REGN10933 (casirivimab) and REGN10987 (imdevimab) in human serum were measured using validated immunoassays which employ streptavidin microplates from Meso Scale Discovery (MSD, Gaithersburg, MD, USA). The methods utilized 2 anti-idiotypic monoclonal antibodies, each specific for either REGN10933 or REGN10987, as the capture antibodies. Captured REGN10933 and REGN10987 were detected using 2 different, non-competing anti-idiotypic monoclonal antibodies, each also specific for either REGN10933 or REGN10987. The bioanalytical methods specifically quantified the concentrations of each anti-SARS-CoV-2 spike monoclonal antibody separately, with no interference from the other antibody. The assay has a lower limit of quantification of 0.156 mg/L for each analyte in the undiluted serum sample.

**Pharmacokinetic Methods**

The pharmacokinetics (PK) analysis population included all participants who received the active study drug (safety population) and who had at least 1 non-missing result following the first dose of the study drug. Participants were analyzed based on actual treatment received. For the sentinel group of Part B, 4 participants were included in the PK analysis set. For the safety group of Part B, 9 participants were included in the PK analysis set. Overall, the PK analysis set was comprised of 13 participants.

Blood samples for measurement of casirivimab and imdevimab concentrations in serum were collected from participants randomized to 1200 mg SC or placebo in the sentinel group of Part B at predose, and on Days 2, 4, 8, 15, 22, 29, 57, 85, 113, 141, 169, 197, and 225. Blood samples for drug concentration also were collected from participants randomized to 1200 mg or placebo in the safety group of Part B at predose, and on Days 29, 57, 113, 169, and 225.

From the above drug concentration blood-sampling schedule, PK parameters were determined by non-compartmental methods (Gibaldi 1982) using Phoenix WinNonlin (version 8.3 Certara, L.P.). Area under the curve in serum from Day 0 to 28 (AUC_0-28_) was determined using the log-linear trapezoidal rule and actual sample collection times. Area under the curve from time zero extrapolated to infinite time (AUC_inf_) was determined as AUC through the last measurable concentration (C_last_) + C_last_/λ_z_, where λ_z_ is the slope of the terminal phase of the concentration-time curve. C_max_ was determined as the maximum observed concentration in serum and t_max_ is the time of maximal concentration in serum. Terminal half-life (t_1/2_) was estimated as 0.693/λ_z_, using actual times.

#### Additional Statistical Methods

All methods described in this section were prespecified (see the Statistical Analysis Plan accompanying this manuscript).

**Hierarchical Testing Sequence**

| **Hierarchy^a^** | **Description** |
| --- | --- |
| Primary | Proportion of participants who subsequently develop signs and symptoms (broad-term) within 14 days of a positive RT-qPCR at baseline or during the 28-day efficacy assessment period^b^ |
| Key secondary | Number of weeks of symptomatic SARS-CoV-2 infection (broad-term) within 14 days of a positive RT-qPCR at baseline or during the efficacy assessment period |
| Key secondary | Number of weeks of high viral load >4 (log_10_ copies/mL) in nasopharyngeal swab samples during the efficacy assessment period |

^a^If statistical significance is established for the primary efficacy endpoint, a hierarchical testing procedure will be applied to the key secondary endpoints at a 2-sided 0.05 significance level. The order of testing sequence for key secondary endpoints is provided in the table.

^b^The primary endpoint was designed to capture symptoms within the incubation period of the disease (ie, up to 14 days after each positive RT-qPCR test) or proximal to a RT-qPCR positive test during the efficacy assessment period. Any participants with symptoms that occurred >14 days after the last positive RT-qPCR were not considered as having a symptomatic infection in the primary analysis. The clause “or during the 28-day efficacy assessment period” was included in the case that there was a Cohort B participant who had a positive RT-qPCR at baseline but next had a negative RT-qPCR (due to sampling error) and then a positive RT-qPCR and then developed symptoms >14 days after dosing but associated with a positive RT-qPCR.

RT-qPCR, quantitative reverse transcription polymerase chain reaction.

**Endpoint Analyses**

In this study, the proportion of households with only a single study participant in the seronegative modified full analysis set B population was more than 80%; therefore, the primary endpoint was analyzed using logistic regression without adjusting for within-household correlation. The model included fixed category effects of treatment group (placebo vs REGEN-COV), region (US vs ex-US), and age (≥12 to <50 years vs ≥50 years).

The key secondary efficacy endpoints were analyzed by Van Elteren tests stratified by region and age.

**Calculation of Number of Weeks**

The duration of symptomatic infection (weeks) is defined as: (the end date of all symptoms minus the date of the first symptom+1)/7. If the symptom is ongoing at the time of the analysis, the end date will be set as the cutoff date for the analysis.

The duration of high viral load (weeks) is calculated by counting the nominal visit weeks during the efficacy assessment period with a high viral load of >4 log_10_ copies/mL.

**Missing Data**

Participants with COVID-19 symptoms who are missing a central lab-determined RT-qPCR test during the efficacy assessment period (eg. are too sick to go to the study site) but have a positive SARS-CoV-2 test from a local lab (eg. in the hospital) are considered to have a symptomatic infection if any of the signs and symptoms of COVID-19 occurred within 14 days of the positive SARS-CoV-2 test observed during the efficacy assessment period.

For virologic endpoints, if NP swab viral load data is missing for a visit, it is not imputed. Only non-missing available NP swab viral load data are used for the analysis of viral load endpoints. Only participants with at least 1 post-baseline viral load data point (in NP swab samples) are included in the analysis.

### Supplementary Results

#### Natural History

We compared the outcome of participants in the placebo group who had no evidence of immunity to SARS-CoV-2 (ie, who were seronegative) and determined that they were at higher risk of developing a symptomatic infection (44/104 [42.3%]) than those with evidence of immunity (ie, who were seropositive; 5/38 [13.2%]). Seronegative participants had higher baseline nasopharyngeal viral loads (**Supplementary Results Figure**) and higher maximum nasopharyngeal viral loads during the efficacy assessment period compared with those who were seropositive (median 5.1 vs 2.6 log_10_ copies/mL, respectively); the average duration of detectable viral RNA per participant was also longer (1.9 vs 1.2 weeks, respectively). Viral load declined more slowly in the placebo group over the first 7 days than was observed in the treatment of symptomatic outpatients, consistent with presentation of these individuals early in the course of infection.(Weinreich et al. 2021: 10.1056/NEJMoa2035002)

**Supplementary Results Figure: Viral Load Over Time in the Placebo Arm by Baseline Serology Status**

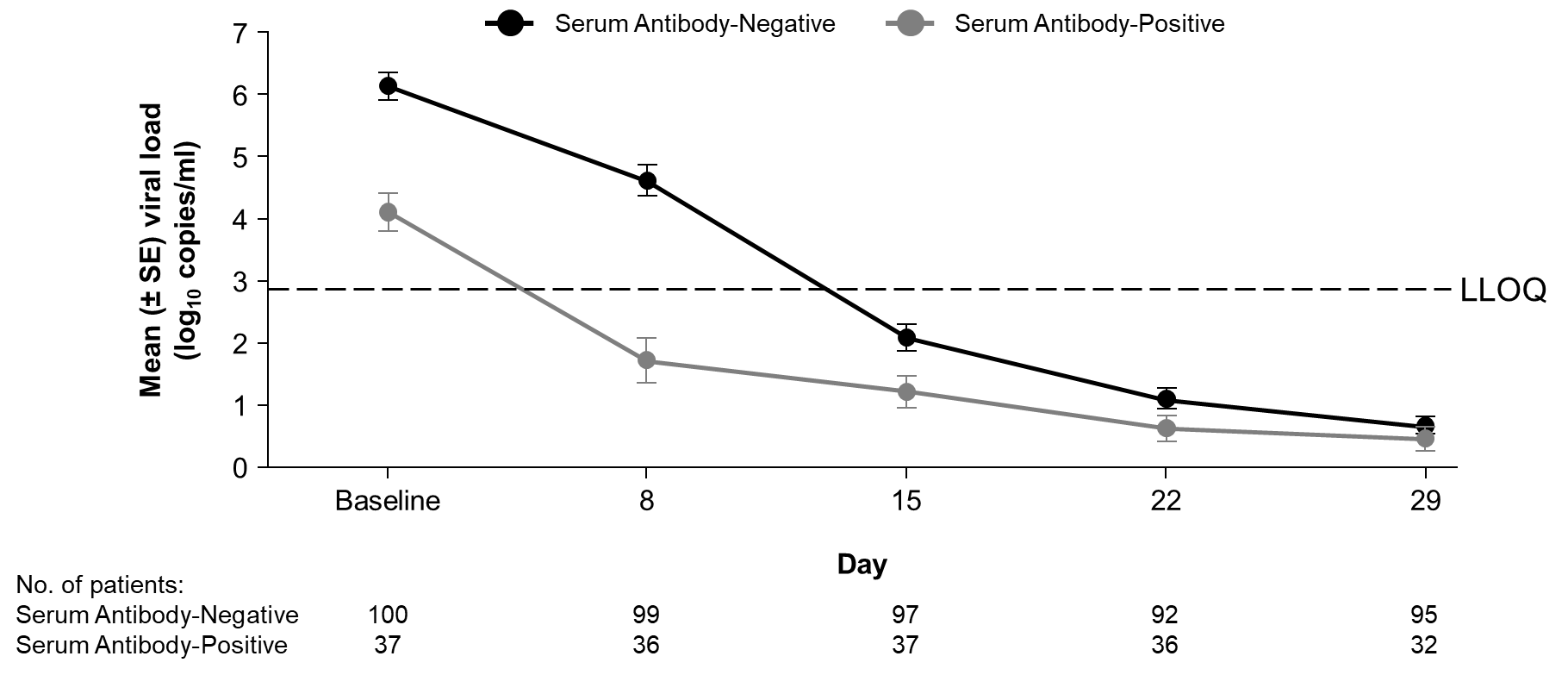

LLOQ, lower limit of quantitation; SE, standard error.

Supplementary Figures
Figure S1. Schematic Overview of the Study Design

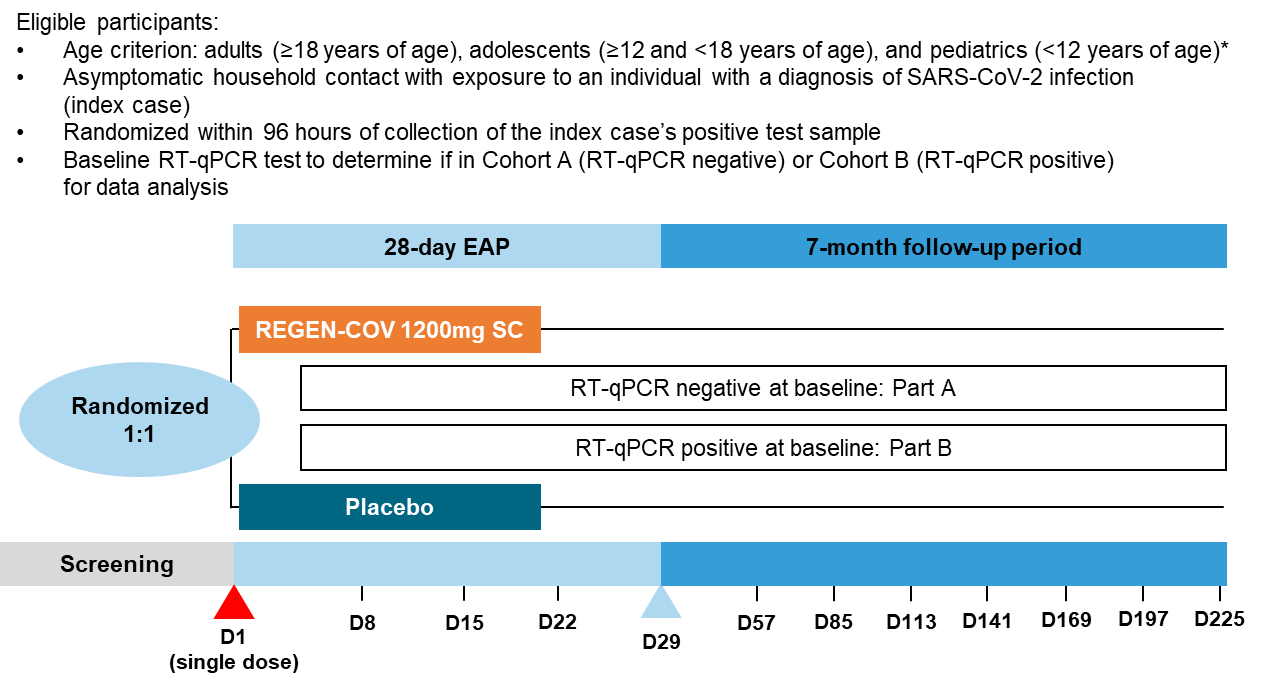

Abbreviations: D, day; EAP, efficacy assessment period; RT-qPCR, quantitative reverse transcription polymerase chain reaction; SC, subcutaneous.
*Pediatric participants are not included in the presented analysis.

Figure S2. Flow Diagram for the Analysis Population

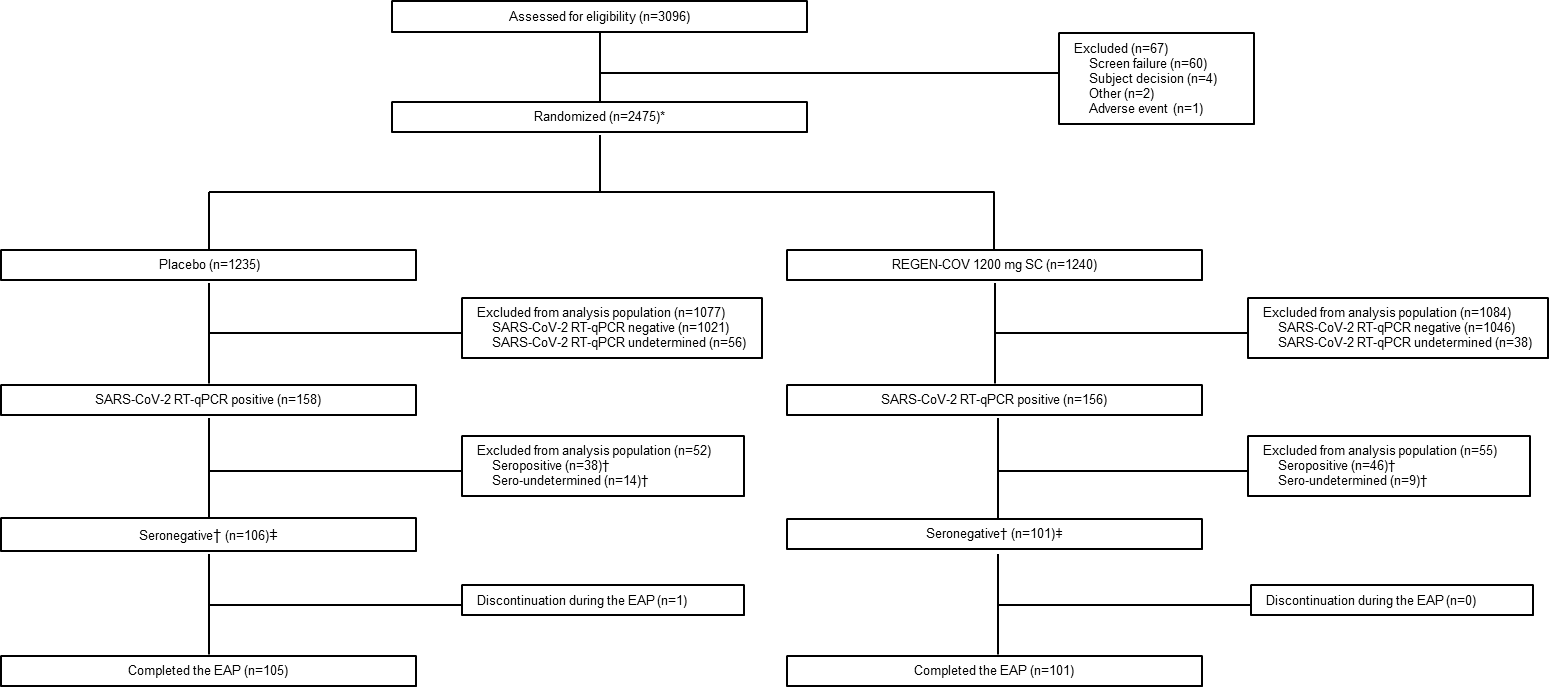

Abbreviations: EAP, efficacy assessment period; mFAS-B, modified full analysis set B; RT-qPCR, quantitative reverse transcription polymerase chain reaction.

*Excludes the 554 participants from a previous interim analysis.

^†^Seronegative: negative result on all available anti-SARS-CoV-2 antibody tests (ie, no evidence of prior infection); seropositive: positive result on 1 or more available anti-SARS-CoV-2 antibody tests; sero-undetermined: borderline result on 1 or more available anti-SARS-CoV-2 antibody tests.

^‡^This is the seronegative mFAS-B population. Among this population, 3 participants (2 in the placebo group and 1 in the REGEN-COV group) were determined post-randomization to have symptomatic disease at baseline and were excluded from the efficacy analyses.

Figure S3. Mean Viral Load in Symptomatic and Asymptomatic SARS-CoV-2 PCR+ Individuals

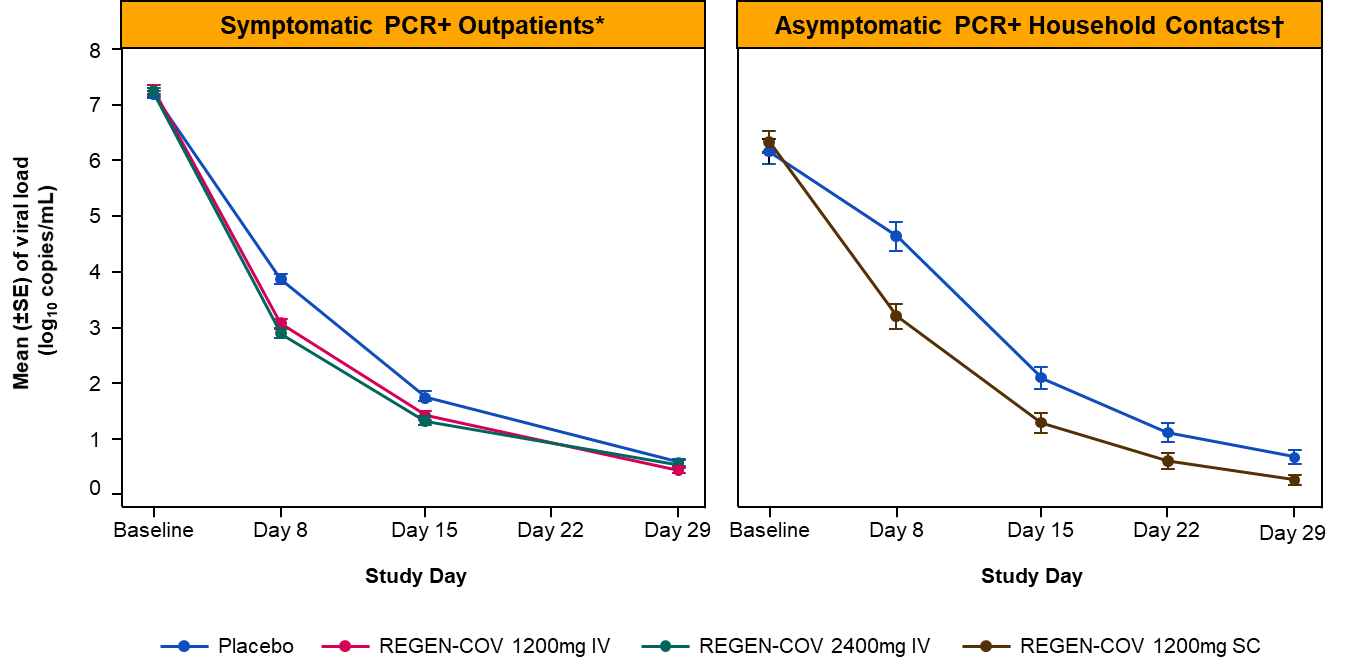

Abbreviations: ANCOVA, analysis of covariance; IV, intravenous(ly); LS, least squares; MMRM, mixed model repeated measures; PCR, polymerase chain reaction; RT-qPCR, reverse transcription quantitative polymerase chain reaction; SC, subcutaneous, SE, standard error.

*Outpatients (Study 2067): symptomatic, RT-qPCR positive, seronegative at baseline; LS mean difference (MMRM) between REGEN-COV and placebo at Day 8: -0.93 log_10_ copies/mL.

^†^Early treatment (Study 2069-B): asymptomatic, RT-qPCR positive, seronegative at baseline; LS mean difference (ANCOVA) between REGEN-COV and placebo at Day 8: -1.5 log_10_ copies/ml.

Figure S4. Mean (+SD) Concentrations of Casirivimab (REGN10933) and Imdevimab (REGN10987) in Serum Over Time (Part B Sentinel Group)

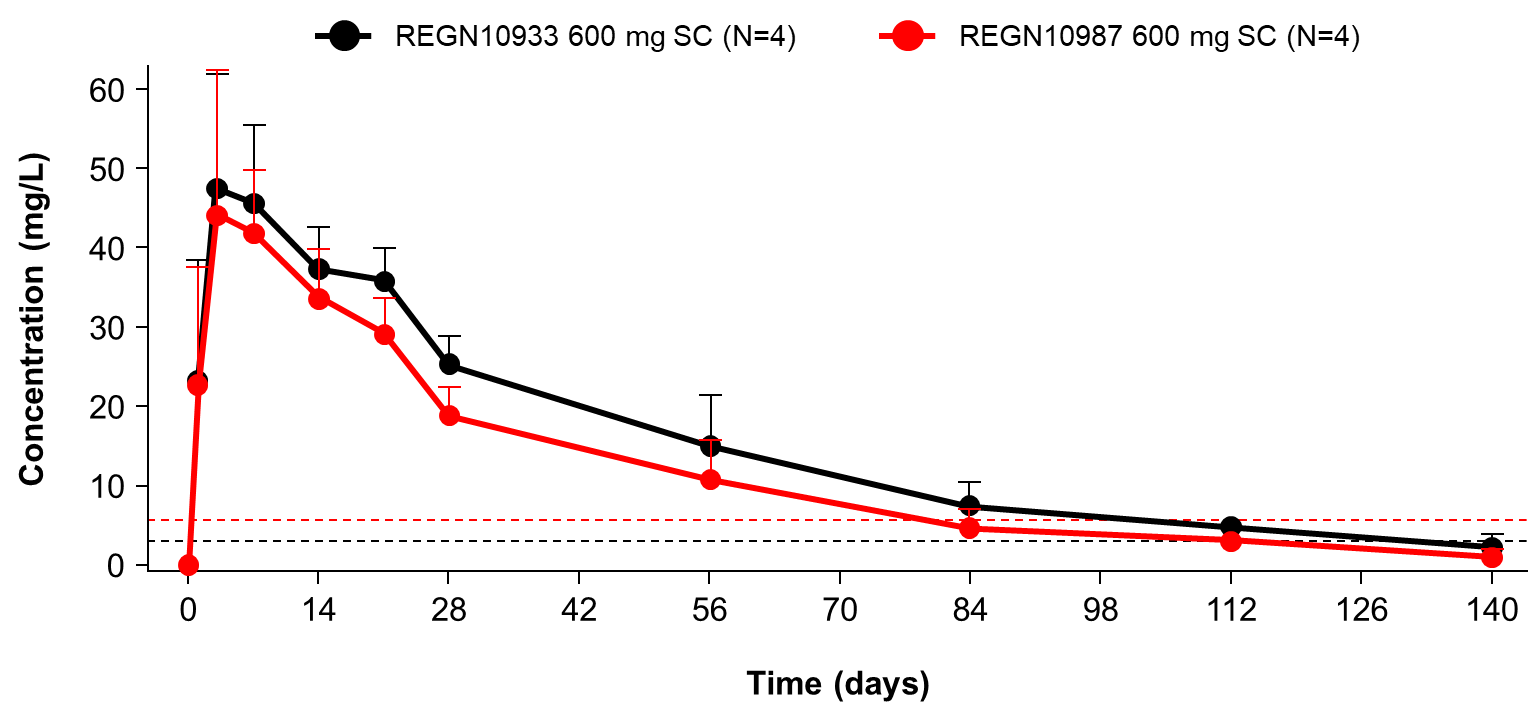

Abbreviations: SC, subcutaneous, SD, standard deviation.

The dashed line denotes 100 * IC90 (delta/B.1.617.2 VOC).

Figure S5. Mean (+SD) Concentrations of Casirivimab (REGN10933) and Imdevimab (REGN10987) in Serum Over Time (Part B Sentinel + Safety Group)

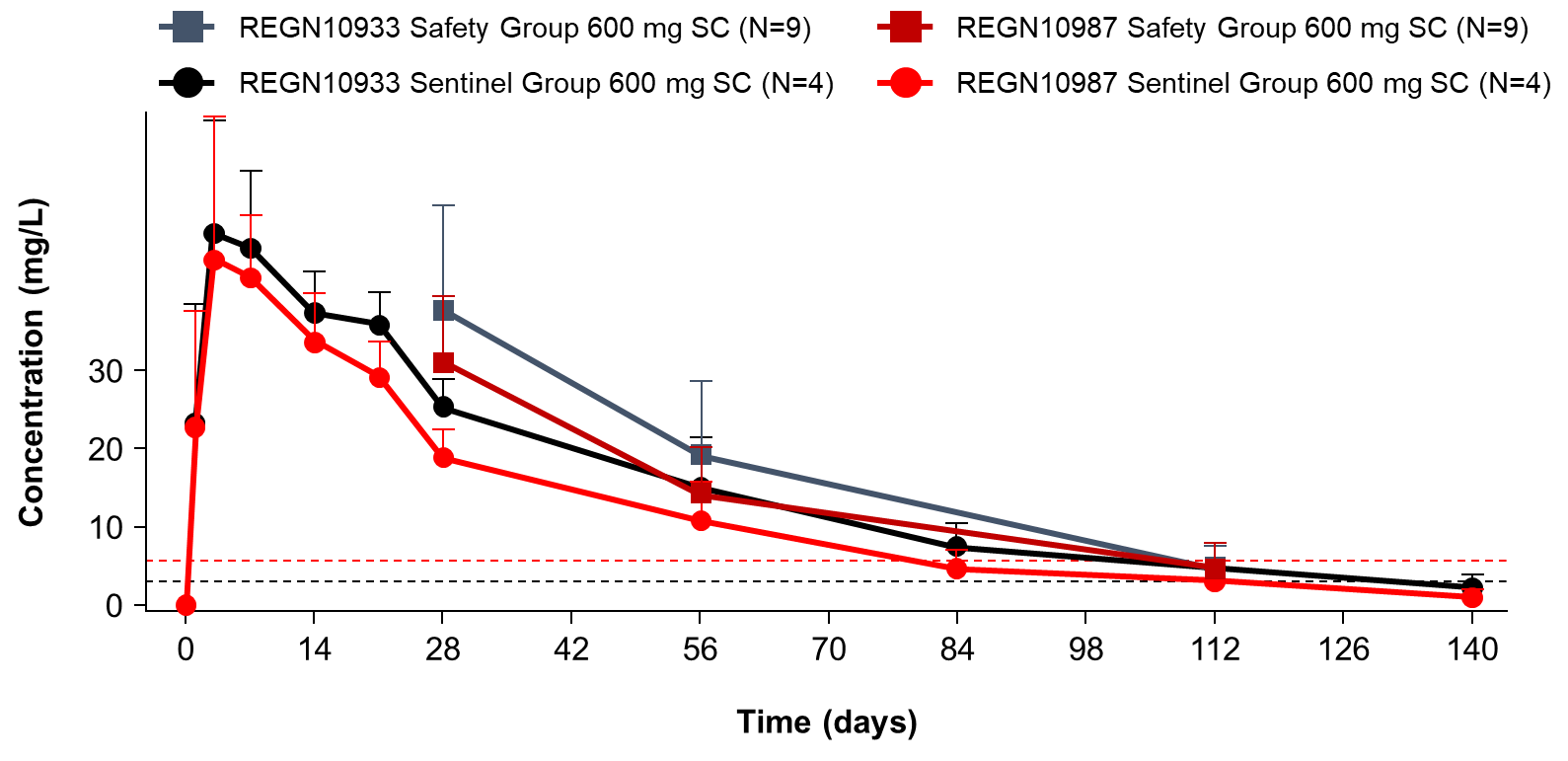

Abbreviations: SC, subcutaneous, SD, standard deviation.

The dashed line denotes 100 * IC90 (delta/B.1.617.2 VOC).

Figure S6. Mean (±SD) Concentrations of Casirivimab and Imdevimab in Serum Over Time (Study 2069 [SC*] vs Study 2067 [IV^†^] or 20145 [IV^‡^])

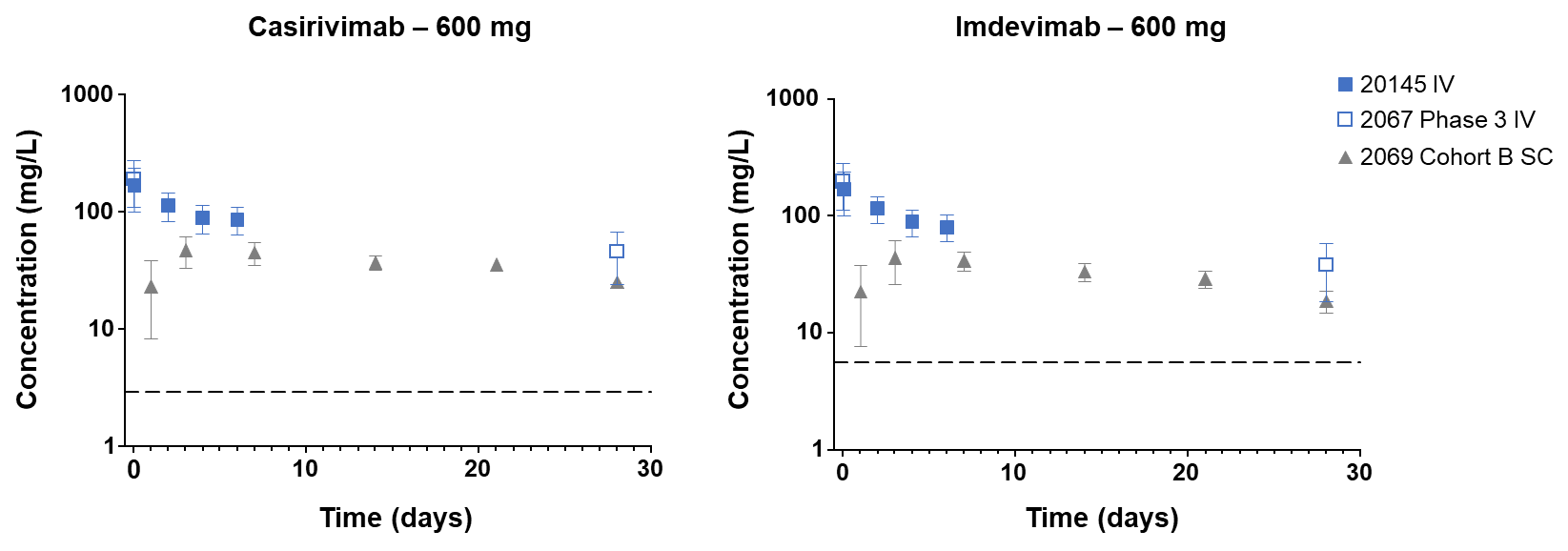

Abbreviations: IV, intravenous; SC, subcutaneous.

*Early treatment (Study 2069-B): asymptomatic, RT-qPCR positive, REGEN-COV 1200 mg SC.

^†^Outpatients (Study 2067): symptomatic, RT-qPCR positive, REGEN-COV 1200 mg IV.

^‡^Outpatients (Study 20145): symptomatic, RT-qPCR positive, REGEN-COV 1200 mg IV.

The dashed line denotes 100 * IC90 (delta VOC).

Supplementary Tables
Table S1. Demographics and Baseline Characteristics (Seropositive)

|  | **Placebo (n = 38)** | **REGEN-COV 1200 mg SC (n = 46)** | **Total (n = 8 4)** |
| --- | --- | --- | --- |
| Age, y |  |  |  |
| Mean (SD) | 39.1 (16.6) | 40.0 (17.9) | 39.6 (17.2) |
| ≥50, no. (%) | 10 (26.3) | 18 (39.1) | 28 (33.3) |
| Male sex, no. (%) | 21 (55.3) | 26 (56.5) | 47 (56.0) |
| Race — no. (%) |  |  |  |
| White | 30 (78.9) | 43 (93.5) | 73 (86.9) |
| Black or African American | 5 (13.2) | 0 | 5 (6.0) |
| Asian | 2 (5.3) | 2 (4.3) | 4 (4.8) |
| American Indian or Alaska Native | 0 | 0 | 0 |
| Other | 1 (2.6) | 1 (2.2) | 2 (2.4) |
| Ethnicity, no. (%) |  |  |  |
| Hispanic or Latino | 17 (44.7) | 26 (56.5) | 43 (51.2) |
| Not Hispanic or Latino | 20 (52.6) | 20 (43.5) | 40 (47.6) |
| Other | 1 (2.6) | 0 | 1 (1.2) |
| Weight, kg, mean (SD) | 79.8 (19.2) | 82.6 (18.8) | 81.4 (18.9) |
| BMI^a^ |  |  |  |
| Mean (SD) | 27.7 (5.6) | 28.5 (5.5) | 28.1 (5.5) |
| >30, no. (%) | 8 (21.1) | 17 (37.0) | 25 (29.8) |
| Total number of households | 36 | 44 | 77 |
| Number of households by size,^b^ no. (%)^c^ |  |  |  |
| 1 | 33 (91.7) | 39 (88.6) | 72 (93.5) |
| 2 | 1 (2.8) | 3 (6.8) | 3 (3.9) |
| 3 | 2 (5.6) | 2 (4.5) | 2 (2.6) |
| >3 | 0 | 0 | 0 |

Abbreviations: BMI, body-mass index; mFAS-B, modified full analysis set-B; SC, subcutaneous; SD, standard deviation.

^a^The BMI is the weight in kilograms divided by the square of the height in meters.

^b^Household size is calculated by counting the seronegative mFAS-B study participants living in the same household.

^c^Percentages based on the total number of households: placebo (n = 36), REGEN-COV (n = 44), and total (n = 77).

Table S2. Proportion of Participants Who Had a Symptomatic RT-qPCR-Confirmed SARS-CoV-2 Infection (Broad-Term) by Period (Seronegative)

| **Period** | **Placebo n/N (%)** | **REGEN-COV 1200 mg SC n/N (%)** | **Relative (Absolute) Risk Reduction, %** | **P-value^a^** |
| --- | --- | --- | --- | --- |
| EAP (all participants) | 44/104 (42.3) | 29/100 (29.0) | 31.5 (13.3) | 0.0380 |
| Day 4 and beyond (after Day 3) | 22/104 (21.2) | 5/100 (5.0) | 76.4 (16.2) | 0.0010^b^ |

Abbreviations: EAP, efficacy assessment period; SC, subcutaneous.

^a^Based on logistic regression model adjusted by region (US vs ex-US) and age group (12 to <50 vs ≥50 years of age).

^b^Nominal.

Table S3. Proportion of Participants Who Had a Symptomatic RT-qPCR-confirmed SARS-CoV-2 Infection by Definition (Seronegative)

|  | **Placebo (n = 104)** | **REGEN-COV 1200 mg SC (n = 100)** |
| --- | --- | --- |
| Proportion of participants who subsequently develop signs and symptoms (broad-term) within 14 days of a positive RT-qPCR at baseline or during the EAP^a^ |  |  |
| n/N (%) | 44/104 (42.3) | 29/100 (29.0) |
| Relative risk reduction, % | - | 31.5 |
| Absolute risk reduction, % | - | 13.3 |
| Odds ratio (95% CI)^b^ | - | 0.54 (0.298, 0.966) |
| P-value^b^ | - | 0.0380 |
| Proportion of participants who subsequently develop signs and symptoms (strict-term) within 14 days of a positive RT-qPCR at baseline or during the EAP |  |  |
| n/N (%) | 20/104 (19.2) | 10/100 (10.0) |
| Relative risk reduction, % | - | 48.0 |
| Absolute risk reduction, % | - | 9.2 |
| Odds ratio (95% CI)^b^ | - | 0.47 (0.207, 1.070) |
| Proportion of participants who subsequently develop signs and symptoms (CDC definition) within 14 days of a positive RT-qPCR at baseline or during the EAP |  |  |
| n/N (%) | 41/104 (39.4) | 27/100 (27.0) |
| Relative risk reduction, % | - | 31.5 |
| Absolute risk reduction, % | - | 12.4 |
| Odds ratio (95% CI)^b^ | - | 0.54 (0.299, 0.989) |

Abbrevaitions: CDC, Centers for Disease Control and Prevention; CI, confidence interval; EAP, efficacy assessment period; RT-qPCR, reverse transcription quantitative polymerase chain reaction; SC, subcutaneous.

^a^Primary endpoint

^b^Based on the logistic regression model adjusted by region (US vs ex-US) and age group (12 to <50 vs ≥50 years of age).

Table S4. Proportion of Participants Who Had a Symptomatic RT-qPCR-confirmed SARS-CoV-2 Infection (Broad-Term) in Those Who Have ≥1 Risk Factor for Severe COVID-19^a^ (Seronegative)

|  | **Placebo (n = 104)** | **REGEN-COV 1200 mg SC (n = 100)** |
| --- | --- | --- |
| Participants with ≥1 risk factor for severe COVID-19, n | 73 | 72 |
| n/N (%) | 31/73 (42.5) | 22/72 (30.6) |
| Relative risk reduction, % | - | 28.0 |
| Absolute risk reduction, % | - | 11.9 |
| Odds ratio (95% CI)^b^ | - | 0.60 (0.284, 1.246) |
| *P*-value | - | 0.1877 |
| Participants with no risk factors for severe COVID-19 (N) | 31 | 28 |
| n/N (%) | 13/31 (41.9) | 7/28 (25.0) |
| Relative risk reduction, % | - | 40.4 |
| Absolute risk reduction, % | - | 16.9 |
| Odds ratio (95% CI)^b^ | - | 0.47 (0.128, 1.593) |
| *P*-value | - | 0.2725 |

Abbreviations: BMI, body mass index; CDC, Centers for Disease Control; CI, confidence interval; EUA, emergency use authorization; RT-qPCR, reverse transcription quantitative polymerase chain reaction; SC, subcutaneous.

^a^Based on a post-hoc analysis of seronegative participants who have ≥1 of the following risk factors for severe COVID-19 (CDC, August 2021): ≥65 years of age, BMI ≥25 kg/m^2^ for adults or ≥85^th^ percentile for adolescents, chronic kidney disease, diabetes, immunosuppressive disease or treatment (ie, immunocompromised state), cardiovascular disease or hypertension, and chronic lung disease.

^b^Based on exact logistic regression model with fixed categorical effects of treatment group.

Table S5: Summary of Viral Load Endpoints (Seronegative)^a^

|  | **Placebo** | **REGEN-COV 1200 mg SC** |
| --- | --- | --- |
| Baseline viral load |  |  |
| n | 97 | 95 |
| Mean (SD) | 6.2 (2.2) | 6.3 (1.9) |
| Change in viral load (log_10_ copies/mL) from baseline to day 8 visits in NP swab samples |  |  |
| n | 97 | 95 |
| Mean (SD) | -1.5 (0.2) | -3.0 (0.2) |
| LS mean difference (SE) vs placebo [95% CI]^b^ | - | -1.5 (0.3) [-2.1 to -0.8] |
| Change in viral load (log_10_ copies/mL) from baseline to day 15 visits in NP swab samples |  |  |
| n | 97 | 95 |
| Mean (SD) | -4.0 (0.2) | -4.9 (0.2) |
| LS mean difference (SE) vs placebo [95% CI]^b^ | - | -0.9 (0.3) [-1.4 to -0.3] |
| Time-weighted average change from baseline in viral load (log_10_ copies/mL) in NP swab samples until the day 22 visit |  |  |
| n | 96 | 95 |
| Mean (SD) | -2.5 (2.3) | -3.7 (1.7) |
| LS mean difference (SE) vs placebo [95% CI]^c^ | - | -0.9 (0.2) [-1.3 to -0.6] |
| Maximum (post-baseline) SARS-CoV-2 RT-qPCR viral load (log_10_ copies/mL) in NP swab samples during the EAP |  |  |
| n | 101 | 98 |
| Mean (SD) | 4.7 (2.6) | 3.3 (2.2) |
| LS mean difference (SE) vs placebo [95% CI]^c^ | - | -1.4 (0.3) [-2.1 to -0.7] |
| Area under the curve in SARS-CoV-2 RT-qPCR viral load (log_10_ copies/mL∙day) in NP swab samples from baseline to the first confirmed negative test |  |  |
| n | 82 | 87 |
| Mean (SD) | 81.7 (50.3) | 56.4 (33.6) |
| LS mean difference (SE) vs placebo [95% CI]^c^ | - | -26.0 (6.0) [-38.0 to -14.1] |

Abbreviations: ANCOVA, analysis of covariance; CI, confidence interval; IQR, interquartile range; LS, least-squares; NP, nasopharyngeal; RT-qPCR, quantitative reverse transcription polymerase chain reaction; SC, subcutaneous; SD, standard deviation; SE, standard error.

^a^Only participants with at least 1 post-baseline viral load data point (in NP swab samples) are included in the analysis.

^b^LS means and SEs taken from the ANCOVA method with the fixed categorical effects of treatment group, age group (age in years: ≥12 to <50 and ≥50), region (US vs ex-US), and the relevant baseline values as covariate as well as treatment-by-covariate interaction.

^c^LS means and SE taken from the ANCOVA method with the fixed categorical effects of treatment group, age group (age in years: ≥12 to <50 and ≥50), region (US vs ex-US), and the relevant baseline values as covariate as well as treatment-by-covariate interaction.

Table S6: Proportion of Participants With a Confirmed Negative SARS-CoV-2 RT-qPCR* During the EAP (Seronegative)

|  | **Placebo (n = 104)** | **REGEN-COV 1200 mg SC (n = 100)** |
| --- | --- | --- |
| Number of participants with a confirmed negative SARS-CoV-2 RT-qPCR^a^ during the EAP |  |  |
| n/N (%) | 70 (67.3) | 86 (86.0) |
| Confirmed negative by day 8 (week 1) |  |  |
| n/N (%) | 13 (12.5) | 23 (23.0) |
| Confirmed negative by day 15 (week 2) |  |  |
| n/N (%) | 37 (35.6) | 56 (56.0) |
| Confirmed negative by day 22 (week 3) |  |  |
| n/N (%) | 57 (54.8) | 78 (78.0) |
| Confirmed negative by day 29 (week 4) |  |  |
| n/N (%) | 70 (67.3) | 86 (86.0) |

Abbreviations: EAP, efficacy assessment period; RT-qPCR, reverse transcriptase quantitative polymerase chain reaction; SC, subcutaneous.

^a^Confirmed negative is defined as 2 consecutive negative SARS-CoV-2 RT-qPCR test results at least 24 hours apart. The time of the first negative is used for analysis.

Table S7. Proportion of Participants Who Had a COVID-19-related Hospitalization or ER Visit (Seronegative)

|  | **Placebo (n = 104)** | **REGEN-COV 1200 mg SC (n = 100)** |
| --- | --- | --- |
| Proportion of participants who had a COVID-19-related hospitalization or ER visit |  |  |
| n/N (%) | 6/104 (5.8) | 0/100 |
| Relative risk reduction | - | 100.0% |
| Absolute risk reduction | - | 5.8% |

Abbreviations: ER, emergency room; SC, subcutaneous.

Table S8: Hospitalization and Hospitalization Outcomes (Seronegative)

|  | **Placebo (n = 104)** | **REGEN-COV 1200 mg SC (n = 100)** |
| --- | --- | --- |
| Proportion of participants who had a hospitalization related to a confirmed SARS-CoV-2 infection up to day 29 |  |  |
| n/N (%) | 3/104 (2.9) | 0/100 |
| Relative risk reduction | - | 100.0% |
| Absolute risk reduction | - | 2.9% |
| Proportion of participants who had a hospitalization related to a confirmed SARS-CoV-2 infection up to day 60 |  |  |
| n/N (%) | 3/104 (2.9) | 0/100 |
| Relative risk reduction | - | 100.0% |
| Absolute risk reduction | - | 2.9% |
| Proportion of participants who were discharged from the hospital among those who were hospitalized due to a confirmed SARS-CoV-2 infection up to day 29 |  |  |
| Hospitalized | 3 | 0 |
| Discharged up to day 29 | 2/3 (66.7) | 0 |
| Discharged up to day 60 | 3/3 (100.0) | 0 |
| Proportion of participants with intubation and mechanical ventilation among those who were hospitalized due to a confirmed SARS-CoV-2 infection |  |  |
| Hospitalized | 3 | 0 |
| With intubation and mechanical ventilation up to day 29 | 0 | 0 |
| With intubation and mechanical ventilation up to day 60 | 0 | 0 |

Abbreviation: SC, subcutaneous.

Table S9. Proportion of Participants Who Had a Symptomatic RT-qPCR-Confirmed SARS-CoV-2 Infection (Broad-Term) by Baseline Serology Status

|  | **Placebo** | **REGEN-COV 1200 mg SC** |
| --- | --- | --- |
| Seronegative^a^ |  |  |
| n/N (%) | 44/104 (42.3) | 29/100 (29.0) |
| Relative risk reduction, % | - | 31.5 |
| Absolute risk reduction, % | - | 13.3 |
| Odds ratio (95% CI)^b^ | - | 0.54 (0.298, 0.966) |
| *P*-value^b^ | - | 0.0380 |
| Seropositive |  |  |
| n/N (%) | 5/38 (13.2) | 4/46 (8.7) |
| Relative risk reduction, % | - | 33.9 |
| Absolute risk reduction, % | - | 4.5 |
| Odds ratio (95% CI)^b^ | - | 0.62 (0.147, 2.587) |
| Seronegative, seropositive, and sero–undetermined |  |  |
| n/N (%) | 53/156 (34.0) | 34/155 (21.9) |
| Relative risk reduction, % | - | 35.4 |
| Absolute risk reduction, % | - | 12.1 |
| Odds ratio (95% CI)^b^ | - | 0.54 (0.325, 0.894) |

Abbrevaitions: CI, confidence interval; SC, subcutaneous.

^a^Primary endpoint.

^b^Based on the logistic regression model adjusted by region (US vs ex-US) and age group (12 to <50 vs ≥50 years of age).

Table S10. Number of Weeks of Symptomatic SARS-CoV-2 Infection (Broad-Term) by Baseline Serology Status

|  | **Placebo** | **REGEN-COV 1200 mg SC** |
| --- | --- | --- |
| Seronegative^a^ |  |  |
| n | 104 | 100 |
| Total, weeks | 170.3 | 89.6 |
| Total per 1000 participants, weeks | 1637.4 | 895.7 |
| Reduction,^b^ % | - | 45.3 |
| *P*-value^c^ | - | 0.0273 |
| Per-symptomatic participant, mean (SD), weeks | 3.9 (4.5) | 3.1 (4.1) |
| Per-participant, mean (SD), weeks | 1.6 (3.5) | 0.9 (2.6) |
| Seropositive |  |  |
| n | 38 | 46 |
| Total, weeks | 30.4 | 9.9 |
| Total per 1000 participants, weeks | 800.8 | 214.3 |
| Reduction ^b^ % | - | 73.2 |
| Per-symptomatic participant, mean (SD), weeks | 6.1 (9.3) | 2.5 (3.4) |
| Per-participant, mean (SD), weeks | 0.8 (3.7) | 0.2 (1.1) |
| Seronegative, seropositive, and sero–undetermined |  |  |
| n | 156 | 155 |
| Total, weeks | 206.3 | 101.7 |
| Total per 1000 participants, weeks | 1322.3 | 656.2 |
| Reduction ^b^ % | - | 50.4 |
| Per-symptomatic participant, mean (SD), weeks | 3.9 (5.0) | 3.0 (3.9) |
| Per-participant, mean (SD), weeks | 1.3 (3.4) | 0.7 (2.2) |

Abbreviations: SC, subcutaneous; SD, standard deviation.

^a^Key secondary efficacy endpoint.

^b^Based on the normalized weeks per 1000 participants.

^c^Based on a stratified Wilcoxon rank sum test (Van Elteren test) with the strata region (US vs ex-US) and age group (age in years: ≥12 to<50 and ≥50).

Table S11. Number of Weeks of High Viral Load (>4 Log_10_ Copies/mL) in NP Swab Samples by Baseline Serology Status

|  | **Placebo** | **REGEN-COV 1200 mg SC** |
| --- | --- | --- |
| Seronegative^a^ |  |  |
| n^b^ | 101 | 98 |
| Total, weeks | 82 | 48 |
| Total per 1000 participants, weeks | 811.9 | 489.8 |
| Reduction,^c^ % | - | 39.7 |
| *P*-value^d^ | - | 0.0010 |
| Per-participant, mean (SD), weeks | 0.8 (0.8) | 0.5 (0.7) |
| Seropositive |  |  |
| n^b^ | 37 | 45 |
| Total, weeks | 6 | 5 |
| Total per 1000 participants, weeks | 162.2 | 111.1 |
| Reduction,^c^ % | - | 31.5 |
| Per-participant, mean (SD), weeks | 0.2 (0.4) | 0.1 (0.4) |
| Seronegative, seropositive, and sero–undetermined |  |  |
| n^b^ | 151 | 152 |
| Total, weeks | 97 | 57 |
| Total per 1000 participants, weeks | 642.4 | 375.0 |
| Reduction,^c^ % | - | 41.6 |
| Per-participant, mean (SD), weeks | 0.6 (0.7) | 0.4 (0.6) |

Abbreviations: NP, nasopharyngeal; SC, subcutaneous; SD, standard deviation.

^a^Key secondary efficacy endpoint.

^b^Only participants with at least 1 post-baseline viral load data point (in NP swab samples) are included in the analysis.

^c^Based on the normalized weeks per 1000 participants.

^d^Based on a stratified Wilcoxon rank sum test (Van Elteren test) with the strata region (US vs ex-US) and age group (age in years: ≥12 to<50 and ≥50).

Table S12. Maximum SARS-CoV-2 RT-qPCR Viral Load (log_10_ copies/mL) in NP Swab Samples by Baseline Serology Status

|  | **Placebo** | **REGEN-COV 1200 mg SC** |
| --- | --- | --- |
| Seronegative^a^ |  |  |
| n | 101 | 98 |
| LS mean (SE) | 4.7 (0.2) | 3.3 (0.2) |
| LS mean difference (SE) vs placebo | - | -1.4 (0.3) |
| 95% CI | - | -2.1, -0.7 |
| Mean (SD) | 4.7 (2.6) | 3.3 (2.2) |
| Median (IQR) | 5.1 (3.4 : 6.3) | 3.6 (2.6 : 4.9) |
| Seropositive^a^ |  |  |
| n | 37 | 45 |
| LS mean (SE) | 2.3 (0.3) | 2.3 (0.2) |
| LS mean difference (SE) vs placebo | - | 0.03 (0.3) |
| 95% CI | - | -0.6, 0.7 |
| Mean (SD) | 2.2 (2.1) | 2.4 (1.6) |
| Median (IQR) | 2.6 (0.0 : 3.7) | 3.0 (0.0 : 3.6) |
| Seronegative, seropositive, and sero–undetermined^a^ |  |  |
| n | 151 | 152 |
| LS mean (SE) | 4.1 (0.2) | 3.0 (0.2) |
| LS mean difference (SE) vs placebo | - | -1.0 (0.3) |
| 95% CI | - | -1.6, -0.5 |
| Mean (SD) | 4.1 (2.6) | 3.0 (2.1) |
| Median (IQR) | 4.0 (2.6 : 6.0) | 3.3 (0.0 : 4.4) |

Abbreviations: ANCOVA, analysis of covariance; CI, confidence interval; IQR, interquartile range; LS, least-squares; NP, nasopharyngeal; RT-qPCR, quantitative reverse transcription polymerase chain reaction; SC, subcutaneous; SD, standard deviation; SE, standard error.

^a^Only participants with at least 1 post-baseline viral load data point (in NP swab samples) are included in the analysis. LS means and SEs taken from the ANCOVA method with the fixed categorical effects of treatment group, age group (age in years: ≥12 to <50 and ≥50), region (US vs ex-US), and the relevant baseline values as covariate as well as treatment-by-covariate interaction.

Table S13. Number of Weeks of RT-qPCR Confirmed SARS-CoV-2 Infection (Regardless of Symptoms) by Baseline Serology Status

|  | **Placebo** | **REGEN-COV 1200 mg SC** |
| --- | --- | --- |
| Seronegative |  |  |
| n | 104 | 100 |
| Total, weeks | 193 | 133 |
| Total per 1000 participants, weeks | 1855.8 | 1330.0 |
| Reduction, % | - | 28.3 |
| Per-participant, mean (SD), weeks | 1.9 (1.3) | 1.3 (1.1) |
| Seropositive |  |  |
| n | 38 | 46 |
| Total, weeks | 47 | 49 |
| Total per 1000 participants, weeks | 1236.8 | 1065.2 |
| Reduction, 5 | - | 13.9 |
| Per-participant, mean (SD), weeks | 1.2 (1.3) | 1.1 (1.0) |
| Seronegative, seropositive, and sero–undetermined |  |  |
| n | 156 | 155 |
| Total, weeks | 268 | 193 |
| Total per 1000 participants, weeks | 1717.9 | 1245.2 |
| Reduction, % | - | 27.5 |
| Per-participant, mean (SD), weeks | 1.7 (1.3) | 1.3 (1.1) |

Abbreviations: RT-qPCR, quantitative reverse transcription polymerase chain reaction; SC, subcutaneous; SD, standard deviation.

Table S14. Treatment-Emergent Adverse Events Occurring in ≥2% of Participants

| **Preferred Term – No. of Participants (%)** | **Placebo (n = 156)** | **REGEN-COV 1200 mg SC (n = 155)** |
| --- | --- | --- |
| COVID-19 | 49 (31.4) | 34 (21.9) |
| Asymptomatic COVID-19^a^ | 12 (7.7) | 7 (4.5) |
| Injection-site reaction | 1 (0.6) | 6 (3.9) |

Abbreviations: AE, adverse event; RT-qPCR, quantitative reverse transcription polymerase chain reaction; SC, subcutaneous; TEAE, treatment-emergent adverse event.

^a^Of these participants, 7 (4.4%) in the placebo group and 5 (3.2%) in the REGEN-COV group were confirmed to have a second positive infection with SARS-CoV-2 after their baseline infection was resolved (ie, 2 negative RT-qPCR test results by central laboratory testing). The remaining 7 participants had a data entry error with the AE start time that resulted in the baseline infection reported as a TEAE; these participants did not have a TEAE of asymptomatic COVID-19.

Table S15. Serious Treatment-Emergent Adverse Events

| **System Organ Class**  **Preferred Term – No. of Participants (%)** | **Placebo (n = 156)** | **REGEN-COV 1200 mg SC (n = 155)** |
| --- | --- | --- |
| Participants with at least 1 serious TEAE | 4 (2.6) | 0 |
| Infections and infestations | 3 (1.9) | 0 |
| COVID-19 | 2 (1.3) | 0 |
| COVID-19 pneumonia | 1 (0.6) | 0 |
| Gastrointestinal disorders | 1 (0.6) | 0 |
| Pancreatitis acute | 1 (0.6) | 0 |

Abbreviations: SC, subcutaneous; TEAE, treatment-emergent adverse event.

Table S16. Summary of PK Parameters for Casirivimab and Imdevimab After a Single 1200 mg Subcutaneous Dose of REGEN-COV in Part B Participants

| **PK Parameter^a^** | **REGN10933 (casirivimab)** | **REGN10987 (imdevimab)** |
| --- | --- | --- |
| C_max_ (mg/L) | 47.5 (12.9) [4] | 46.1 (13.8) [4] |
| t_max_ (day)^b^ | 7.50 (4.00, 9.00) [4] | 7.50 [4.00, 9.00] [4] |
| AUC_0-28_ (mg●day/L) | 953 (213) [4] | 840 (183) [4] |
| AUC_inf_ (mg●day/L)^c^ | 2104 (515) [4] | 1609 (419) [4] |
| C_28_ (mg/L)^c–e^ | 33.5 (12.3) [9] | 26.9 (9.12) [9] |
| Half-life (day) | 30.2 (5.31) [4] | 26.5 (5.31) [4] |

^a^Mean (SD) [N].

^b^Median (range) [N].

^c^Value reported for subjects with %AUC_inf_ extrapolated <20%.

^d^Observed concentration 28 days after dosing, ie, on day 29, as defined in the protocol.

^e^Value represents subjects in the sentinel and safety groups.
